# Automating Clinical Phenotyping Using Natural Language Processing: An Application for Crohn’s Disease

**DOI:** 10.1101/2023.10.16.23297099

**Authors:** Linea Schmidt, Susanne Ibing, Florian Borchert, Julian Hugo, Allison A. Marshall, Jellyana Peraza, Judy H. Cho, Erwin P. Böttinger, Bernhard Y. Renard, Ryan C. Ungaro

## Abstract

Real-world studies based on electronic health records often require manual chart review to derive patients’ clinical phenotypes, a labor-intensive task with limited scalability. Here, we developed and compared computable phenotyping based on rules using the spaCy framework and a Large Language Model (LLM), GPT-4, for disease behavior and age at diagnosis of Crohn’s disease patients. We are the first to describe computable phenotyping algorithms using clinical texts for these complex tasks with previously described inter-annotator agreements between 0.54 and 0.98. The data comprised clinical notes and radiology reports from 584 Mount Sinai Health System patients. Overall, we observed similar or better performance using GPT-4 compared to the rules. On a note-level, the F1 score was at least 0.90 for disease behavior and 0.82 for age at diagnosis. We could not find statistical evidence for a difference to the performance of human experts on this task. Our findings underline the potential of LLMs for computable phenotyping.

**Graphical Abstract:** 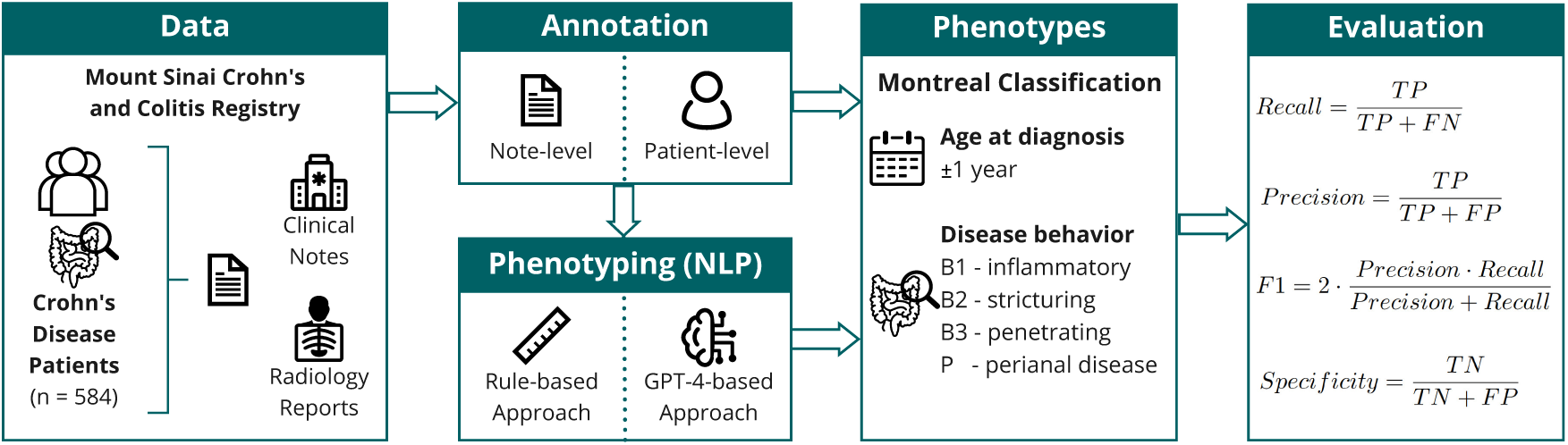

## 1 Introduction

Computable clinical phenotyping, the automatic grouping of patients according to their medical history captured in Electronic Health Records (EHR), has been the focus of numerous studies in the past decades [1], [2]. For these approaches, manual chart review is limited to developing and validating the algorithms, therefore enabling clinical research at scale. Large consortia such as Electronic Medical Records and Genomics (eMERGE) have invested in resources such as the Phenotype Knowledge Base, which focuses on developing, sharing, and validating computable phenotyping algorithms [3].

These algorithms can broadly be divided into two main categories: rule-based and Machine Learning (ML)-based approaches. The choice between these approaches relies on factors like the availability of data and the complexity of the task at hand [4]. Rule-based techniques utilize predefined rules or criteria, typically requiring expertise in the relevant medical domain. In contrast, ML-based methods employ algorithms to recognize data patterns corresponding to different phenotypes.

Often, phenotypes of patient subgroups are insufficiently captured in the structured EHR, requiring analysis of clinical narrative text and the use of Natural Language Processing (NLP) to derive these phenotypes [5]. In their benchmark paper, Moldwin *et al.* demonstrated, amongst others, for digestive diseases, that the incorporation of unstructured data outperforms models that are only based on structured EHR [6]. While ML-based approaches may be able to identify unknown patterns using vast amounts of data, a rule-based approach provides increased transparency compared to ML approaches, particularly when applying Large Language Models (LLMs).

As Ananthakrishnan *et al.* demonstrated, one example of phenotypes that cannot be reliably extracted from solely structured EHR are clinical subgroups of Crohn’s Disease (CD) [7]. CD, one of the main types of Inflammatory Bowel Disease (IBD), is an immune-mediated disease marked by recurrent episodes of chronic inflammation of the gastrointestinal (GI) tract. The disease is characterized by significant heterogeneity in disease course and treatment response [8]. In a recent publication, an expert consensus of the European Crohn’s and Colitis Organization (ECCO) discussed core outcomes and outcome measures relevant to be reported in IBD studies based on real-world data, such as EHR [9]. Even though randomized studies are considered gold-standard, studies based on real-world data are of interest to derive complementary evidence. The data is essential as it enables longitudinal analysis of rich clinical data beyond the time and patient population constraints of clinical trials [10]. For instance, studies based on real-world data can mitigate the fact that many patients with severe disease courses are not included in clinical trials [11]. According to the ECCO expert group, when reporting or studying disease complications, it is recommended to consider the disease phenotype, such as the presence of strictures or fistulas, as core outcomes. The Montreal Classification was recommended as an outcome measure [9] and is used to group CD patients considering three phenotype categories: *age at diagnosis*, *disease behavior*, and *disease location* (Table 1) [12], [13].

**Table 1:** Montreal Classification for Crohn’s Disease patients according to Silverberg, Satsangi, Ahmad, *et al.* [12]. *^†^*“p” is added to B1–B3 when concomitant perianal disease is present. *^∗^*L4 is a modifier that can be added to L1–L3 when concomitant upper gastrointestinal disease is present.

| Category | Classification | Definition |
| --- | --- | --- |
| Age at Diagnosis | A1 | Below 16 years |
|  | A2 | Between 17 and 40 years |
|  | A3 | Above 40 years |
| Disease Behavior | B1 | Non-stricturing and non-penetrating |
|  | B2 | Stricturing |
|  | B3 | Penetrating |
|  | p<sup>†</sup> | Perianal disease modifier |
| Disease Location | L1 | Ileal |
|  | L2 | Colonic |
|  | L3 | Ileocolonic |
|  | L4* | Isolated upper disease |

Disease behavior comprises different disease complications of CD, for instance, strictures (B2) or fistulas and abscesses (B3). Strictures are luminal narrowings in any part of the GI tract. They are developed due to chronic inflammation of the mucosa, resulting in excessive repairs of the area of inflammation and, eventually, the mixture of inflamed and scarred tissue [14]. Fistulae are abnormal connections between different parts of the GI tract, between the GI tract and other organs, or between the GI tract and the skin. They can develop due to chronic inflammation and damage to the intestinal wall [8], [15]. In the context of CD, an abscess is a localized accumulation of pus that can develop due to inflammation or infection, due to complicated and active disease [16]. Perianal disease is regarded as a disease modifier that can co-occur with any other disease phenotype (B1-B3): Any penetrating or stricturing disease complication in the perianal region is considered perianal disease. Disease behavior is not a static classification category since disease complications can be gained during the course of the disease [17]. The nuanced and often complex descriptions of the behavioral phenotype in clinical text include descriptions of symptoms and treatment responses, as well as the progression of the disease (Figure 1). Since phenotype data are stored mainly in clinical narrative text, a patient’s disease behavior is usually extracted by manual chart review [18]–[20].

**Figure 1:**
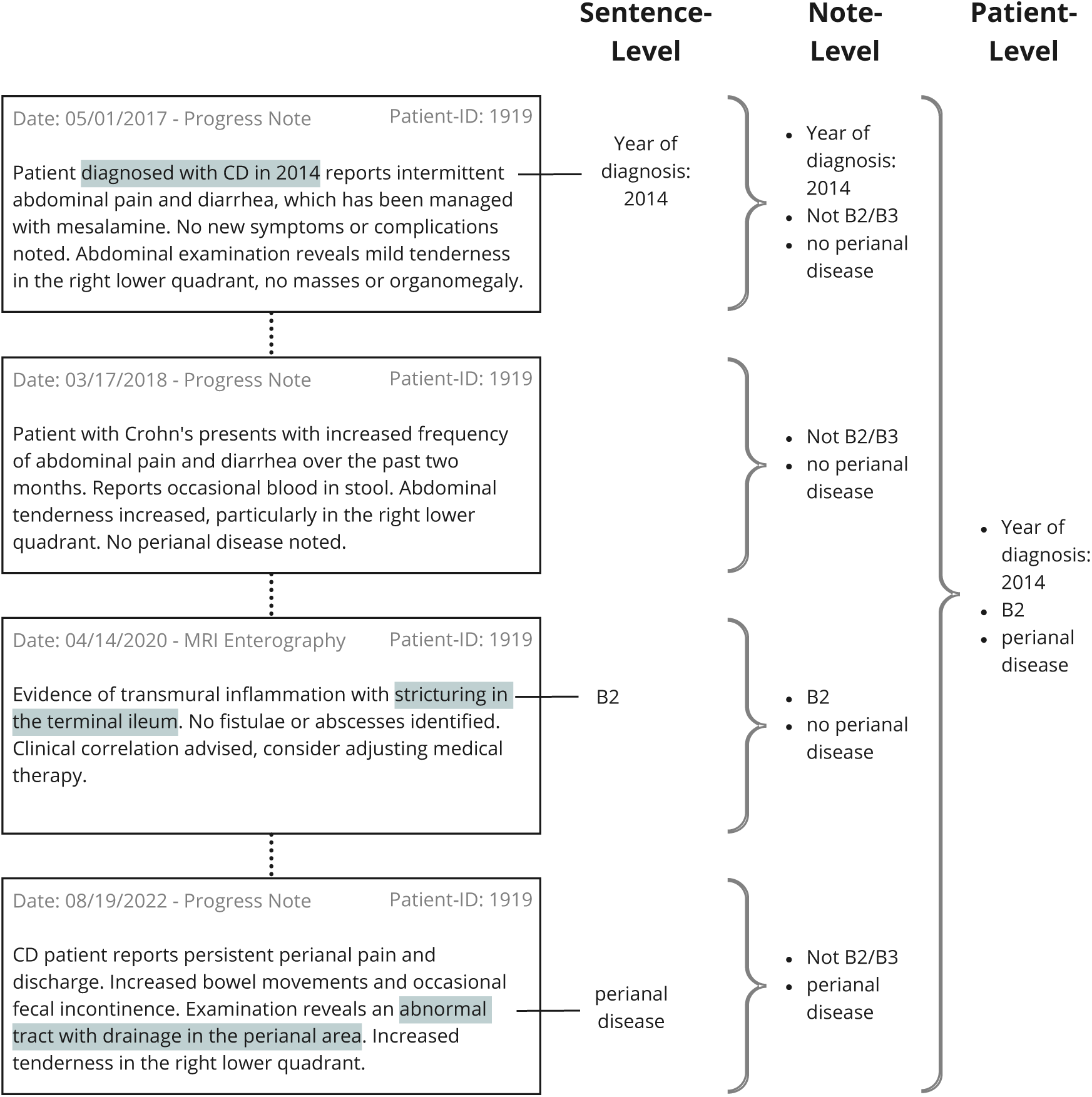
Sentence-, note-, and patient-level labeling for an example patient.

Age at diagnosis refers to the age at initial CD diagnosis, and disease location to the intestinal region of inflammation. Next to disease behavior, age at diagnosis is an essential clinical component for CD clinical care and study cohort characterization, as it allows the deduction of disease duration, a prognostic factor for treatment response with biologics [21]. Automated phenotyping based on NLP techniques, including information from clinical notes, could facilitate patient classification on a large scale with minimal manual labeling required. For instance, Stidham *et al.* recently demonstrated the successful extraction of extraintestinal manifestations in IBD patients using a rule-based NLP approach [22].

In this work, we describe the development of two novel, sentence-based labeled datasets, including annotations of disease phenotype and age at diagnosis in clinical notes of CD patients. We used these data to develop and evaluate rule-based phenotyping algorithms and compare them with an in-context learning approach using a GPT-4 model from OpenAI. The established pipeline facilitates the large-scale labeling of previously unannotated clinical narrative text.

## 2 Materials and Methods

### 2.1 Data Collection and Preprocessing

Clinical notes from the EHR of the Mount Sinai Data Warehouse (MSDW) [23] were acquired via the Artificial Intelligence Ready Mount Sinai (AIR·MS) platform. This dataset was further enriched with reports from the radiology department, allowing the inclusion of Computed Tomography (CT) and Magnetic Resonance Imaging (MRI) reports. 792 CD patients from the Mount Sinai Crohn’s and Colitis Registry (MSCCR) were considered for this study, as previously described [18]. This cohort was selected due to already existing annotations on a patient-level. We preprocessed the available clinical notes and radiology reports by removing irrelevant note types (e.g., telephone encounters and patient instructions) and texts that did not contain a CD-related context (Figure 2). 584 of the 792 patients had at least one non-empty clinical note available after filtering. The clinical text dates range from February 1940 (first clinical text) to May 2023 (latest clinical text). For the annotation and extraction of the age at diagnosis, we additionally filtered notes for regular expressions containing key expressions such as “diagnosed”, “Crohn’s […] since”, or “age at”^1^. To allow for further granularity, we split all clinical texts into sentences (Figure 2).

**Figure 2:**
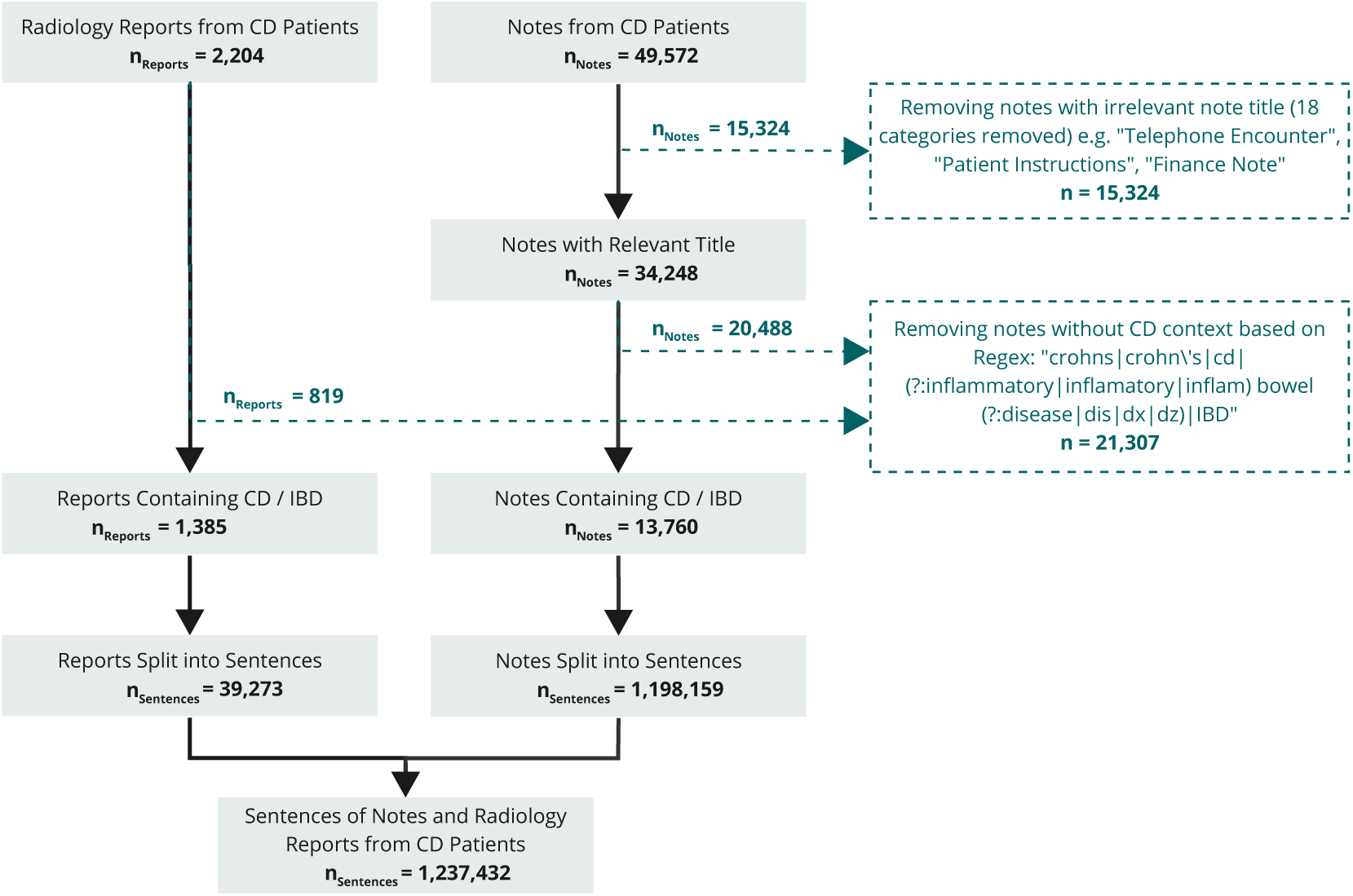
Data sources and preprocessing steps. After extracting all available clinical notes from CD patients in MSCCR up until two weeks after the date of initial endoscopy and biopsy for sample collection for the study, all notes with irrelevant tiles were removed. Subsequently, from the available clinical notes and radiology reports, only disease-relevant texts were further processed by splitting them into sentences.

### 2.2 Annotation and Dataset Creation

For disease behavior, annotation guidelines were based on Montreal classification annotation procedures from the COMPASS-IBD study and the Ocean State Crohn’s and Colitis Area Registry (OSCCAR) data dictionaries [19], [20]. Two annotators, an internal medicine resident, and an IBD researcher, labeled 200 notes on a sentence-level (Figure 3). An agreement sample of 50 notes (5,543 sentences) was used to ascertain the Inter-Annotator Agreement (IAA), which was measured using Cohen’s kappa statistics [24]. After resolving disagreements, a curated dataset was created and used as a test set. We additionally labeled a development set consisting of 200 clinical texts, labeled by non-experts. Rules were exclusively developed using this development dataset and evaluated on previously unseen test data. To allow an evaluation of the disease behavior on the patient level, we used a labeled subset from MSCCR. The data comprised 134 labeled patients with available clinical texts until their first endoscopy within the MSCCR study.

**Figure 3:**
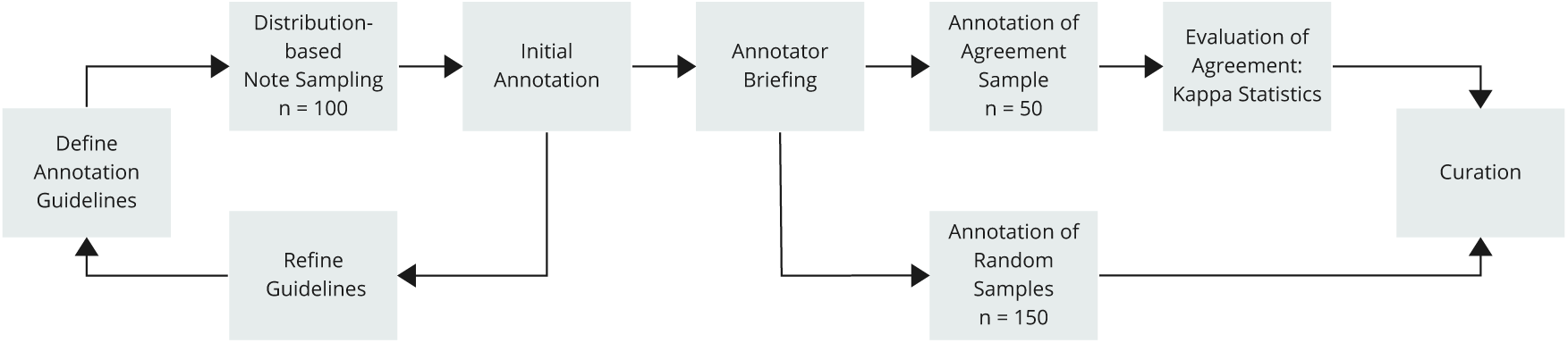
Labeling process. The process from building annotation guidelines to a final annotated and curated dataset for CD disease complications containing 150 clinical notes and 50 radiology reports.

The annotation of the age at diagnosis was conducted by the IBD researcher, labeling 200 additional clinical texts on a sentence level based on three categories: age at diagnosis, diagnosis year, and disease duration. Notably, the ground-truth age at diagnosis was calculated using patients’ birth years and the note dates. After curation, this dataset was split into a testing and a development sets. The original labels from MSCCR patients with available clinical texts were used for a patient-level evaluation.

### 2.3 Disease Behavior Phenotyping

Two primary methods for disease behavior phenotyping were adopted: a rule-based algorithm and an in-context learning approach.

#### 2.3.1 Rule-based Approach

The rule-based approach leverages spaCy [25](for syntactic analysis and pattern matching), scispaCy [26] (for concept extraction), and medspaCy (for negation detection) [27]. A custom spaCy component, BehavioralPhenoCategorizer, is constructed for phenotype extraction. The *en core sci md* scispaCy model is the cornerstone for syntactic analyses and named entity recognition. After preprocessing, abbreviation detection, and Unified Medical Language System (UMLS) linking using a curated subset of UMLS Metathesaurus codes, patterns were established to detect specific CD behavioral phenotypes. The development process utilized spaCy’s Matcher class to design patterns that describe token sequences for accurate disease phenotyping of CD. Multiple patterns were crafted: for specific phenotype complications, direct string-level matches, UMLS linkages, and two additional patterns addressing medical conjectures (uncertainty matcher) and explicit exclusions (exclusion matcher). These patterns were refined to differentiate complications like B2/B3 from perianal disease through UMLS linking and token-level regular expression-like patterns.

In clinical texts, the presence and absence of medical conditions are often described, making effective negation detection crucial. For behavioral phenotyping in CD, two strategies were adopted: one leveraging medspaCy, a rule-based approach that identifies negation patterns and uses dependency parsing to determine negated entities, and the other utilizing a Transformer-based Clinical Assertion and Negation Classification BERT model [28]. For the latter, we deployed the pre-trained *bvanaken/clinical-assertion-negation-bert* model from the Hugging Face Hub^2^ that detects entity absence with a probability score, considering spans as negated if they surpass a 0.5 threshold.

The BehavioralPhenoCategorizer processes each document in stages: initial categorization using UMLS matching, pattern application to detect matches, followed by exclusion checks based on direct string matching of terms such as “no” or “not” and the results of the chosen negation detection method. In case of a CD complication match, a context window of up to seven tokens is scanned for uncertainty or exclusion patterns. If the match is not determined to be negated but still linked to B2 or B3 classifications, proximity to mentioning the perianal region is checked, leading to potential phenotype reassignment. Phenotypes are stored and aggregated at different levels (patient, note, or sentence), with the most severe phenotype following the order B1*<*B2*<*B3. The phenotyping performance is assessed if the input data contains labeled ground-truth information (Figure 4).

**Figure 4:**
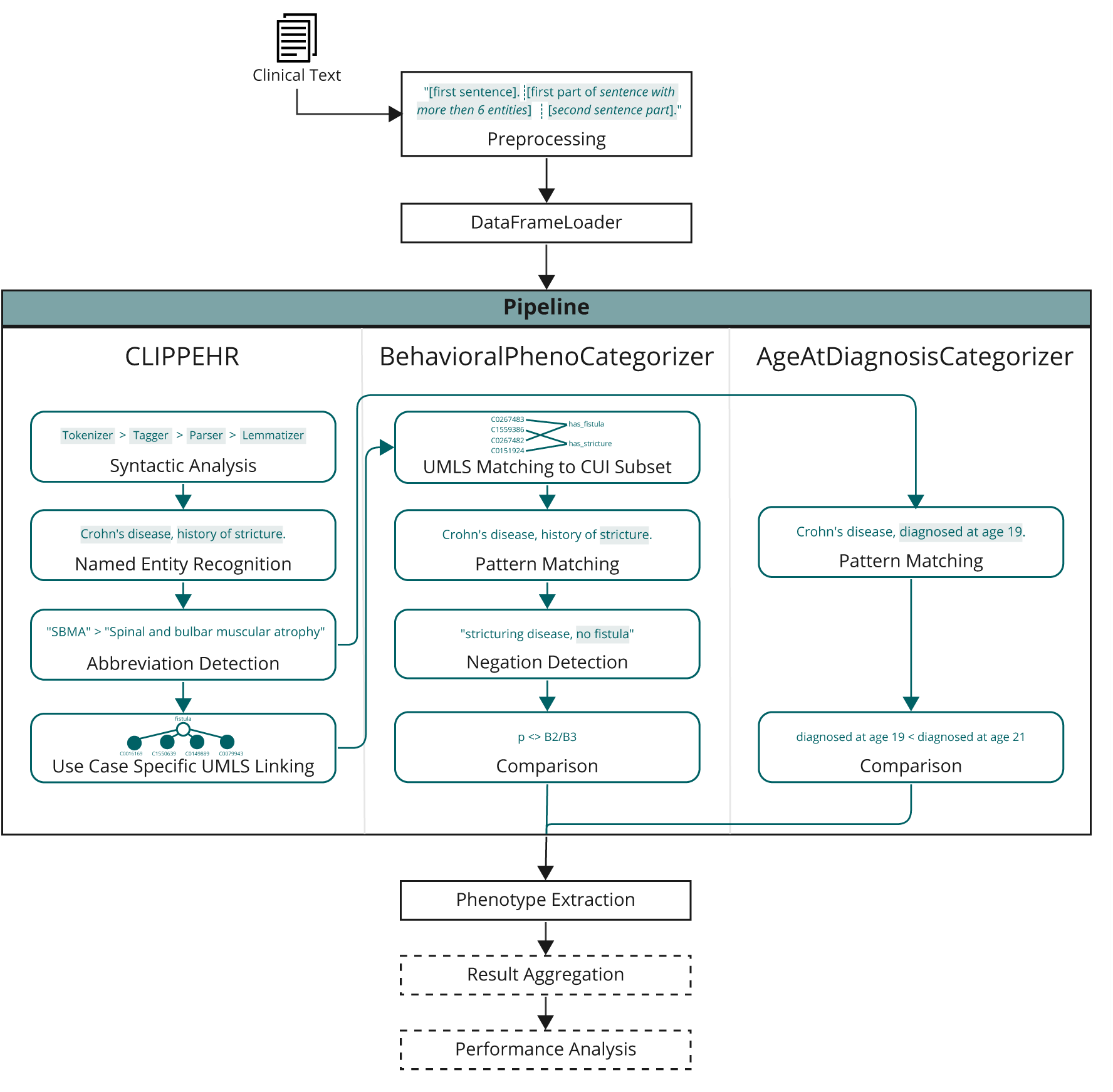
Rule-based phenotyping algorithm. The phenotyping process starts with clinical texts from radiology and clinical notes as input. The spaCy pipeline contains, on the one hand, elements defined by CLIPPEHR and, on the other hand, the newly developed BehavioralPhenoCategorizer or the AgeAtDiagnosisCategorizer. After the phenotype extraction, result aggregation and performance analysis are optional. Evaluation is only conducted if ground-truth labels are available.

#### 2.3.2 In-context Learning

We deployed the Azure OpenAI GPT-4 model (1106-Preview) to generate note-level structured output on disease behavior phenotypes [29]. For the prompt, we added the slightly shortened annotation guidelines of the COMPASS-IBD study and three randomly selected examples, including outputs from the development dataset, an approach considered as fewshot prompting (Table A1). The model was used off-the-shelf without any fine-tuning. We implemented a function call to pre-define and structure the output of the GPT-4 model. To make the model results more deterministic, we set the temperature parameter of the model to 0. Besides that, the OpenAI default parameters were used.

### 2.4 Age at Diagnosis Phenotyping

Similar to the disease behavior, a custom spaCy component, AgeAtDiagnosisClassifier, was engineered to determine the age at diagnosis. Through a series of pattern matching, textual spans indicating age at diagnosis (e.g., “diagnosed with 8 years old”), disease duration (e.g., “CD since 10 years”), or diagnosis year (e.g., “CD diagnosed in 2002”) were recognized. Subsequent analysis determined the age at diagnosis based on these matches, the patient’s year of birth, and the date of the clinical text. Of note, the identified year of birth includes an error margin of ±1 year since exact dates are not frequently mentioned in clinical notes. True Positives (TP) corresponded to accurately identified ages at diagnosis (within ±1-year) for performance metrics. True Negatives (TN) were accurate identifications where age information was absent, False Positives (FP) were defined as wrongly identified age at diagnosis labels, and False Negatives (FN) represented overlooked labeled instances.

We used the same prompt and model parameters as for disease behavior phenotyping for the in-context learning approach (Table A1, section 2.3.2).

### 2.5 Algorithm Performance Evaluation

The phenotyping algorithms were evaluated on note- and patient-level using the annotated test datasets. The evaluation was conducted based on recall, precision, specificity, and F1 score. During the development of the disease behavior rule-based phenotyping algorithm using the separate development dataset, our primary metric of interest was recall, given the importance of the sensitive identification of positive instances. For age at diagnosis, on the other hand, we prioritized precision to focus on the accurate extraction of the information for downstream tasks. Maintaining a balanced precision, F1 score, and specificity were set as secondary aims.

## 3 Results

### 3.1 Curation of the Disease Behavior and Age at Diagnosis Datasets

To evaluate the performance of the disease behavior phenotyping algorithms, we created a newly annotated dataset comprising 150 clinical notes and 50 radiology reports, with a total of 15,390 sentences (Table A2).

50 of these clinical notes were labeled by two different annotators. The quality of the labeling process was determined through Cohen’s kappa agreement scores. On sentence-level, we observed an overall IAA score of 0.85 (Not B2/B3: 0.83; B2/B3: 0.84; perianal disease: 0.87; Table A3). Evaluation on note-level increased the overall IAA to 0.90. These results indicate a good consensus among annotators [30], underlining the robustness and appropriateness of the labeled data as ground truth for subsequent phenotyping algorithm evaluations.

After the annotators found a consensus for all disagreement instances, in the finalized, curated test set, approximately 1% of clinical note sentences and 3.6% of radiology report sentences got a B2 or B3 label assigned. 0.8% of clinical note sentences and 2.1% of radiology report sentences were annotated with perianal disease (Table A2).

For age at diagnosis, we labeled a total of 80 randomly selected clinical texts comprising 79 clinical notes and one radiology report, amounting to a total of 12,293 unique sentences (Table A4). Most commonly, in 60 % of the texts, the year of diagnosis was given, followed by the age of diagnosis (23 % of texts) and disease duration (10 % of texts).

### 3.2 Effective Disease Behavior Extraction on Note-Level with Rules and GPT-4

While we only used full note texts as input for GPT-4, we ran the rule-based approach at sentence-level, further aggregating the results for note- or patient-level evaluation. To optimize our spaCy pipeline, we conducted a series of experiments with varied settings regarding rule types and negation detection options on the development set at the sentence level. Concerning the differentiation of rule types, we observed that a synergistic approach combining UMLS matching rules with rules for direct string matching was superior in its performance compared to applying either of the rule types alone. Notably, the exclusive employment of string matching exhibited superior results compared to relying solely on UMLS matching across clinical notes and radiology reports (Figure A1). For negation detection, we analyzed the number of false positives and false negative disease complications using either the medspaCy negation detection component, the Large Language Model (LLM) Negation Classifier, or no negation detection. The LLM Negation Classifier performed superior to the other two options, manifesting the lowest incidence of false negatives while preserving a substantial number of accurately identified instances (Figure A2, Figure A3).

Using either the rule-based or LLM-based approach, the automated behavioral phenotyping based on clinical notes yielded high recall values on note-level, ranging from 0.92 - 1.00, depending on the phenotype (Table 2). The identification of no existing disease complications was particularly successful, with recall and precision values above 0.94 for both the GPT-4 and rule-based approach. Overall, the two approaches performed very similarly on note-level. Also, when extracting the disease complication categories individually, we were able to identify all instances of perianal disease and B2 (Table A5). Nonetheless, we improved the results by aggregating B2 and B3 into one joint disease complication category compared to separately phenotyping B2 and B3. The calculated Cohen’s kappa agreement scores between the labels from the annotated consensus dataset and the labels derived from the rules or GPT-4 are on average between 0.83 and 0.86 (Table A3). These scores are comparable and statistically not inferior to the IAA between the human annotators (Table A6), underlining the high performance of the computational approaches.

**Table 2:** Performance of the rule-based phenotyping algorithms and in-context learning using a GPT-4 model to extract disease behavior on note-level using the newly annotated test datasets comprised of 150 clinical notes. Stricturing and penetrating complications were combined into one “B2/B3” category. Rules AND GPT-4: Only instances with same results from rules and GPT-4 are considered; Rules OR GPT-4: disease complication was considered if labeled by either rules or GPT-4.

| Phenotype | Model | Recall | Precision | F1 score | Specificity |
| --- | --- | --- | --- | --- | --- |
| **Not B2/B3** | Rules | 0.94 | 1.00 | 0.97 | 0.84 |
|  | GPT-4 | 0.95 | 0.98 | 0.96 | 0.86 |
|  | Rules AND GPT-4 | 0.98 | 1.00 | 0.99 | 0.94 |
|  | Rule OR GPT-4 | 0.90 | 1.00 | 0.95 | 0.78 |
| **B2/B3** | Rules | 1.00 | 0.84 | 0.92 | 1.00 |
|  | GPT-4 | 0.95 | 0.86 | 0.90 | 0.98 |
|  | Rules AND GPT-4 | 1.00 | 0.94 | 0.97 | 1.00 |
|  | Rule OR GPT-4 | 1.00 | 0.78 | 0.87 | 1.00 |
| **Perianal - yes** | Rules | 1.00 | 0.86 | 0.93 | 0.86 |
|  | GPT-4 | 0.92 | 0.92 | 0.92 | 0.92 |
|  | Rules AND GPT-4 | 1.00 | 1.00 | 1.00 | 1.00 |
|  | Rule OR GPT-4 | 1.00 | 0.81 | 0.89 | 0.81 |
| **Perianal - no** | Rules | 0.97 | 1.00 | 0.98 | 1.00 |
|  | GPT-4 | 0.98 | 0.98 | 0.98 | 0.98 |
|  | Rules AND GPT-4 | 1.00 | 1.00 | 1.00 | 1.00 |
|  | Rule OR GPT-4 | 0.95 | 1.00 | 0.98 | 1.00 |

Using radiology reports, the model performance values dropped to 0.64 - 1.00, with less sensitive identification, particularly of B3 and B2 (Table A7). The underlying reason may be the incorrect identification of B2 or B3 as perianal disease. The precision values and F1 scores indicate over-classification, resulting in false positive disease complication labels.

We additionally tested the performance of combining outputs from the GPT-4 and rule-based approaches (Table 2). When considering disease complications, if at least labeled by one of the two approaches (Rules OR GPT-4), we improved recall for detection of disease complications (1.00). However, we lost precision (0.81 for perianal disease, 0.78 for structuring or penetrating disease). Only considering instances with the same labels based on the rules and GPT-4 yielded the highest performance metrics with balanced recall and precision, with F1 scores between 0.97 and 1.00. The overlap between labels derived from the GPT-4 model and the rules for perianal disease was at 95%, for *Not B2/B3* and *B2/B3* at 90%.

### 3.3 Joint Evaluation of Rules and GPT-4 Enables Chart-Review Prioritization

For 134 patients in the MSCCR study, we extracted disease phenotype at study enrollment through manual chart review, considering all clinical information up until this point. Compared to the note-level analysis, using the rule-based approach, we achieved a recall value of 0.71 and a precision of 0.48 for detecting any complication (B2 or B3). For the detection of perianal disease, the recall was 0.85 and precision 0.56, decreased compared to the note-level analysis (Table 3).

**Table 3:** Performance of rule-based phenotyping algorithms and in-context learning using a GPT4-model to extract disease behavior on patient-level. 134 patients of the MSCCR cohort had available information on the behavioral disease phenotype through manual chart review. Stricturing and penetrating complications were combined into one “B2/B3” category. Rules AND GPT-4: Only instances with same results from rules and GPT-4 are considered; Rules OR GPT-4: disease complication was considered if labeled by either rules or GPT-4.

| Phenotype | Model | Recall | Precision | F1 score | Specificity |
| --- | --- | --- | --- | --- | --- |
| **Not B2/B3** | Rules | 0.65 | 0.83 | 0.73 | 0.48 |
|  | GPT-4 | 0.86 | 0.84 | 0.85 | 0.68 |
|  | Rules AND GPT-4 | 0.87 | 0.83 | 0.85 | 0.72 |
|  | Rule OR GPT-4 | 0.64 | 0.83 | 0.72 | 0.48 |
| **B2/B3** | Rules | 0.71 | 0.48 | 0.58 | 0.83 |
|  | GPT-4 | 0.64 | 0.68 | 0.66 | 0.84 |
|  | Rules AND GPT-4 | 0.66 | 0.72 | 0.69 | 0.83 |
|  | Rule OR GPT-4 | 0.71 | 0.48 | 0.57 | 0.83 |
| **Perianal - yes** | Rules | 0.85 | 0.56 | 0.68 | 0.56 |
|  | GPT-4 | 0.78 | 0.58 | 0.67 | 0.58 |
|  | Rules AND GPT-4 | 0.84 | 0.68 | 0.75 | 0.68 |
|  | Rule OR GPT-4 | 0.85 | 0.50 | 0.63 | 0.50 |
| **Perianal - no** | Rules | 0.83 | 0.96 | 0.89 | 0.96 |
|  | GPT-4 | 0.86 | 0.94 | 0.90 | 0.94 |
|  | Rules AND GPT-4 | 0.89 | 0.95 | 0.92 | 0.95 |
|  | Rule OR GPT-4 | 0.79 | 0.95 | 0.86 | 0.95 |

Also for the GPT-4 based approach, the performance dropped considerably compared to note-level analysis. Similar to the note-level results, aggregating the data into one disease complication category improved results (Table A8).

Similar to the note-level evaluation, we applied ensemble methods based on the combined output from the two approaches, either by focusing only on patients with the same labels (77% overlap for Not B2/B3 and B2/B3 and 89% overlap for perianal disease) or by considering a disease complication if detected by either of the approaches. Only considering overlapping labels yielded more balance between recall and precision, with F1 scores of 0.69 (B2/B3) and 0.75 (perianal disease).

The GPT-4 model alone performed well at identifying the non-existence of disease complications (no B2/B3 and no perianal disease), with an F1 score of 0.85 and 0.90, respectively. This reflects a reduction of false positives compared to the rule-based approach. The F1 score was further improved for perianal disease by 0.02 using the overlap ensemble approach.

We evaluated the count of the note-level labels for each patient sub-group to understand better whether a count-based cut-off may help to further stratify patients for chart review. Patients labeled as B2/B3 had a significantly higher number of notes in their records classified as B2/B3 compared to patients without any labeled complication (Figure A4). Similarly, patients diagnosed with perianal disease have a noticeably higher frequency of notes relating to this condition (Figure A5). However, for a considerable number of patients (rules: 15, GPT-4: 12) with known disease complications, this information was not extracted from or noted in any of the clinical notes. Therefore, stratification based on note count with extracted disease complication did not remain a suitable option.

Of note, for the annotation process of the patient-level ground-truth labels, as the primary clinical information system was used, the basis of underlying data differed from the information available for the automated phenotyping.

### 3.4 GPT-4 Outperforms Rule-based Approach in Age at Diagnosis Extraction

We evaluated the performance of the age at diagnosis extraction through the rules and GPT-4 model on note- and patient-level (Table 4). Using the rule-based approach, we observed balanced performance metrics on note-level, exemplified by a recall of 0.78 and a precision of 0.87. The GPT-4 model outperformed the rule-based model with a recall of 1.00 and a precision value of 0.87. The correlation between the extracted ages at diagnosis was very high with 0.98 for GPT-4 (Figure A6). While also showing a higher correlation compared to the rule-based approach (*R* = 0.88), the GPT-4 model was also able to label all notes that included information on the age at diagnosis (64 out of 80). In contrast, the rules only recognized 79% of the notes. Aggregating the results from GPT-4 and the rule-based approach by calculating the mean or only including instances where the extracted age at diagnosis was the same between rules and GPT-4 did not considerably improve overall results.

**Table 4:** Performance of rule-based phenotyping algorithms and in-context learning using a GPT4-model to extract age at diagnosis on note- and patient-level. 80 clinical notes were newly annotated for evaluation of this task. Patient-level performance evaluation was conducted using 458 MSCCR patients with available clinical narrative texts and existing age at diagnosis labels of the cohort. NA values are based on the fact, that all patients have an annotated age at diagnosis, resulting in 0 true negatives.

| Phenotype | Evaluation | Model | Recall | Precision | F1 score | Specificity |
| --- | --- | --- | --- | --- | --- | --- |
| **Age at diagnosis** | Note-level | Rule-based | 0.78 | 0.87 | 0.82 | 0.68 |
|  |  | GPT-4 | 1.00 | 0.87 | 0.93 | 0.53 |
|  | Patient-level | Rule-based | 0.73 | 0.70 | 0.71 | NA |
|  |  | GPT-4 | 1.00 | 0.59 | 0.74 | NA |

On patient-level, a similar balance of the performance measures was achieved using the rule-based approach, with lower overall performance values compared to the note-level, with F1 scores of 0.71 and 0.82, respectively. Conversely, the metrics from GPT-4 were not as balanced, with a recall of 1.00 and a precision of 0.59, but overall a higher F1 score (0.74) compared to the rule-based approach. The correlation of GPT-4 and the rule-based age at diagnosis with the manually labeled values was high (GPT-4: *R* = 0.84; Rules: *R* = 0.81), but reduced compared to the note-level (Figure A7). Only considering patients with the same labels yielded an R value of 0.86, but compared to the GPT-4 based approach, a reduction of extracted labels by 34%.

The underlying data of the 458 MSCCR patients for evaluation on patient-level was not specifically scanned for availability of the information in scope within the written clinical text. Therefore, the reduced performance of the model on patient-level may be explained by the limited data availability.

## 4 Discussion

In the current study, we demonstrate the feasibility of automatically extracting clinical information on CD disease behavior and age at diagnosis from EHR using a rule-based and LLM-based approach. To our knowledge, we are the first to describe NLP-based phenotyping for the stated tasks and to conduct a comparison between rules and the GPT-4 model. We observed that GPT-4 performed as well or better than the rule-based approaches, suggesting that large general-purpose language models may provide a more efficient and better-performing method to automated disease phenotyping in IBD.

The IAA of manual chart review of the described phenotypes differed between previous studies, with Cohen’s kappa values between 0.54 and 0.79 for disease behavior and between 0.67 and 0.98 for age at diagnosis [31]–[33]. These findings underscore the complexity of both tasks, in particular for the extraction of disease behavior. To evaluate our algorithms, we created two datasets annotated on sentence-level. For disease behavior, we included two annotators, and their IAA, on average 0.85 to 0.90, surpassed the kappa statistics stated in previous studies. This may be due to differences in annotators training or abstraction rules compared to other studies. Of note, the sentence- or note-level annotation agreements are difficult to be compared with patient-level annotations.

Shrestha *et al.* described in their work the identifications of disease phenotypes by using International Classification of Diseases (ICD)-codes of the Swedish National Patient Register [34]. For our use case, working with ICD-codes was not a suitable option: Given the fragmented nature of the US healthcare system [35], coded information in EHR data is not sufficiently reliable to extract the complex clinical information [7]. Nevertheless, the published study by Shrestha *et al.* provides a baseline for computable phenotyping with which we can compare our patient-level results. Their reported recall values lie between 0.62 for B2/B3, 0.75 for B1, and 0.81 for perianal disease, and on average 0.94 for the different phenotype groups of age at diagnosis. Our results are overall comparable when only considering patients with identical labels using the rules and GPT-4, even exceeding the results reported based on the ICD codes from the Swedish Patient register. Furthermore, our reported performances are similar to those described for other tasks in the literature, such as the extraction of extraintestinal manifestations [22].

The presented rule-based models offer systematic and transparent reasoning and are thus potentially the preferred support for labeling tasks in a clinical setting, especially when no baseline for the stated problem exists. The newer general-purpose LLM, GPT-4, achieved very similar results as the rule-based approach when extracting the patients’ disease behavior. The model’s capacity to digest and consider the context of one clinical note as a whole is of interest for context-dependent phenotyping tasks. In particular, for age at diagnosis, the recall of the GPT-4 model with 1.00 on note- and patient-level was considerably higher than for the rule-based approach with 0.78 and 0.73, respectively. While on patient-level this increased value is accompanied by a reduction of precision by 0.11, the extraction of time or basic demographic information may be an easier task than the extraction of medical domain-specific information for a general LLM such as GPT-4 [36]. In addition, an important consideration is the time effort required to develop the different approaches: the estimated time effort to develop the rules for disease behavior, including the development of the task-specific UMLS subset for matching, was about two months of full-time work and for the rules for age at diagnosis extraction an additional 2 weeks. The implementation and development of the prompt for GPT-4 took about a day. Our approach to incorporating the annotation guidelines into the prompt, in addition to randomly selected examples from the development set, provided a quick and successful solution for prompt engineering.

Our models for disease behavior classification performed superior on clinical notes compared to radiology reports (Table 2). Radiology reports were underrepresented in our test and development sets, which may have influenced performance. Furthermore, they may miss important elements of the patients’ clinical data, such as doctor office visit findings or the patient history. Additionally, these reports typically feature longer sentences, intricate language structures, fewer spelling errors, and frequent suggestions, exclusions, and negations, highlighting the need for broader representation and potentially different processing strategies in future studies.

For a more in-depth understanding of false classifications on a patient level, we analyzed five falsely positive and falsely negative classified patients for B2/B3 and perianal disease. False positive instances for B2/B3 mainly came from describing similar complications in other disease contexts (e.g., carotid stenosis), complex sentence structures leading to errors in negation detection, and confusion with perianal disease labels. Instances misclassified as perianal disease are suspected to be partly wrong-labeled. In one instance, the negation detection was not sophisticated enough to catch the negation in the given sentence structure. There was no clear description of the phenotype in the clinical texts for the patients with false negative labels of B2/B3 and perianal disease. Challenges for the age at diagnosis extraction were mainly caused by the varied representations of dates in the data, coupled with the task of unambiguously linking a date occurrence to the diagnosis of CD.

While our study shows promising results, we acknowledge certain limitations. Foremost, due to the fragmentation of clinical data into multiple IT systems, we did not have access to endoscopy or pathology reports for our study. Addition of these data types may improve future studies. Furthermore, our phenotyping pipeline is based only on information captured in clinical text and does not incorporate structured EHR data. With this underlying difference of utilized data for computable phenotyping compared to manual chart review, and without checking whether the information we are looking for is, in fact, captured in our underlying data, the patient-level evaluation has to be regarded with caution. Second, our models were only evaluated on internal clinical texts from Mount Sinai Health System (MSHS). This poses a potential limitation, as the rule-based algorithms might be particularly tuned to language idiosyncrasies specific to physicians within the Mount Sinai health system or the reporting conventions typical of this institution. Therefore, future external validations efforts will be needed to finalize understanding of the models’ generalizability in other clinical settings, which may have different linguistic nuances or documentation practices.

In conclusion, GPT-4 performed at least comparable and, in some cases, better than the rule-based approach for CD disease phenotyping, with little effort involved. Therefore, we anticipate that LLMs will be increasingly deployed for phenotyping tasks. Despite the reduced performance of the described phenotyping algorithms on patient-level, we are confident that our work can, in the future, contribute to studies based on large patient cohorts. Prioritization of patients for chart review may be one future application in order to accelerate the labeling process and improve patient quality. Here, patients labeled as non-penetrating, non-stricturing and non-perianal disease by the two approaches may be reviewed with less time spent, as the automated phenotyping performed very well in these cases. Furthermore, for patients with automatically labeled disease complications, the additional information on the date of the clinical note where the complication is spotted may enable a more targeted chart review. Our approach can serve as a strong baseline for such future developments. For future work, the comparison with domain-specific LLM such as MEDITRON or BioMistral for complex domain-specific tasks such as extraction of disease behavior, and for tasks requiring less domain-expertise, such as extraction of age at diagnosis, may be of interest [37], [38].

## Data Availability

The data are not publicly available as it contains protected health information.

## Author contributions

Linea Schmidt (Data Curation: Equal; Methodology: Equal; Formal analysis: Lead; Writing - Original Draft: Equal; Writing – review & editing: Equal)

Susanne Ibing (Conceptualization: Equal; Data Curation: Equal; Methodology: Equal; Formal analysis: Supporting; Writing - Original Draft: Equal; Writing – review & editing: Equal)

Florian Borchert (Conceptualization: Equal; Methodology: Equal; Formal analysis: Supporting; Supervision: Equal; Writing – review & editing: Equal)

Julian Hugo (Data Curation: Supporting; Writing – review & editing: Equal)

Allison A. Marshall (Data Curation: Equal; Writing – review & editing: Equal)

Jellyana Peraza (Data Curation: Equal; Writing – review & editing: Equal)

Judy H. Cho (Supervision: Supporting; Writing – review & editing: Equal)

Erwin P. Böttinger (Supervision: Supporting; Writing – review & editing: Equal)

Bernhard Y. Renard (Supervision: Supporting; Writing – review & editing: Equal)

Ryan C. Ungaro (Conceptualization: Equal; Supervision: Equal; Writing – review & editing: Equal)

## Data and code availability

The data used for this study contain protected health information (PHI) and cannot be shared publicly due to patient privacy reasons. The underlying code for this study is available on github and can be accessed via this link: https://gitlab.com/dacs-hpi/cd-nlp-phenotyping.

## Acknowledgements

We thank all participants of the Mount Sinai Crohn’s and Colitis Registry. Further, we thank Eugenia Alleva, Jan Philipp Sachs, and Gabriele Campanella for their support and fruitful discussions, Danielle Cohen, Alexandra Deutschenbaur, Narges Ghaedi Bardeh, Daniel Jühling, Lisa Koeritz, Larissa Röhrig, Marco Schaarschmidt, Jonathan Wilke, and Paul Wullenweber for their development of the CLIPPEHR pipeline, Manbir Singh, Herve DiBello, and Lewis Lo for IT and data access support, and Marco Pereanez and the Mount Sinai Imaging Research Warehouse for providing the radiology reports. This work is supported in part through the use of the research platform AI-Ready Mount Sinai (AIR*·*MS) and the expertise provided by the team at the Hasso Plattner Institute for Digital Health at Mount Sinai (HPI*·*MS). This work was supported by the Office of Research Infrastructure of the National Institutes of Health under award number S10OD026880. This work was supported in part through the computational and data resources and staff expertise provided by Scientific Computing and Data at the Icahn School of Medicine at Mount Sinai and supported by the Clinical and Translational Science Awards (CTSA) grant UL1TR004419 from the National Center for Advancing Translational Sciences. This work was supported by the NIH K23 Career Development Award K23KD111995-01A1 and NIH R03 DK132440-01A1 (RCU), U01DK062422 (JHC), the Sanford J. Grossman Charitable Trust (JHC), R01DK123758 (JHC), the Joachim-Herz Foundation (SI), the HPI Research School on Data Science and Engineering (LS), the Friedrich Wingert Foundation (LS), and the Hasso Plattner Foundation (HPF).

## Supplementary Tables

**Table A1:**
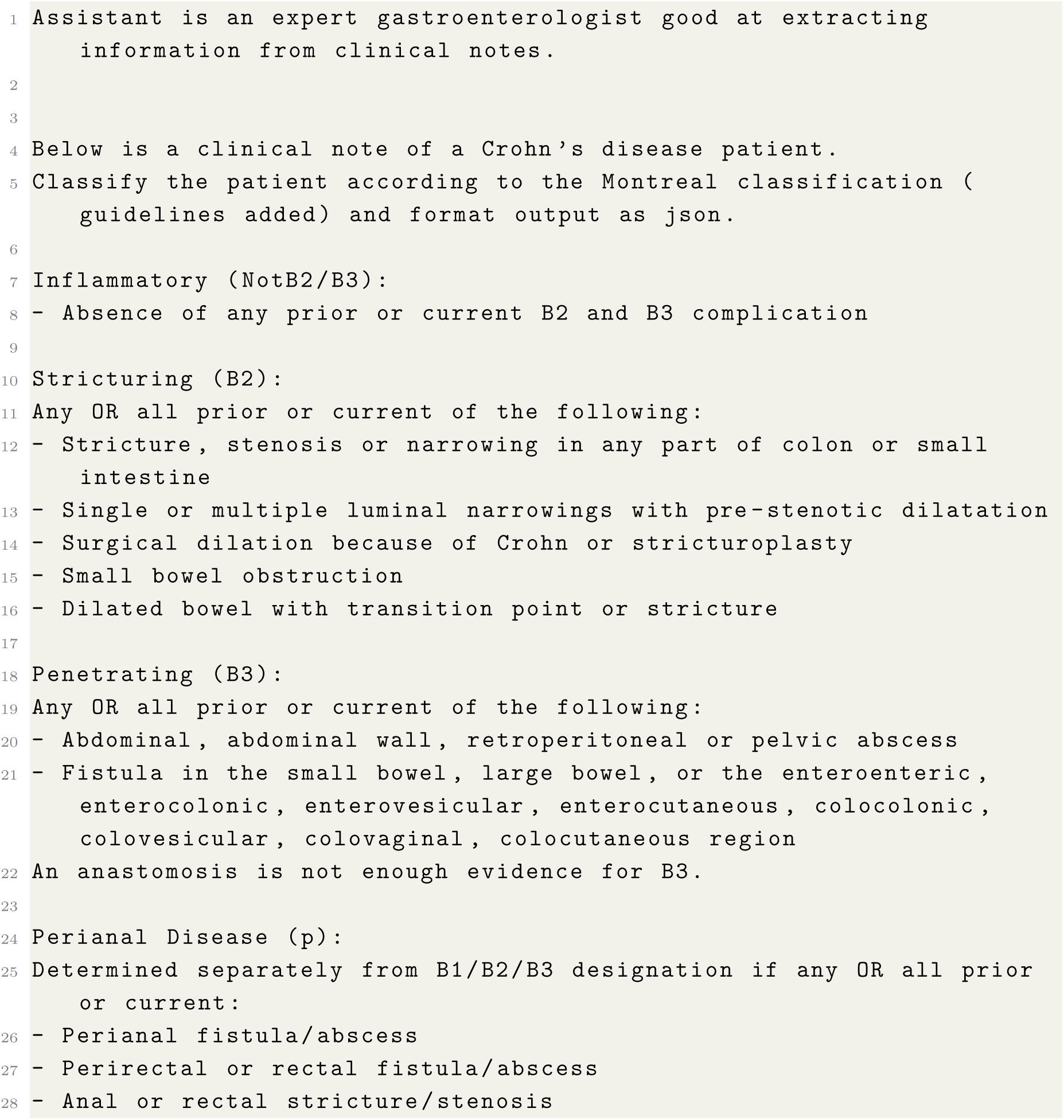
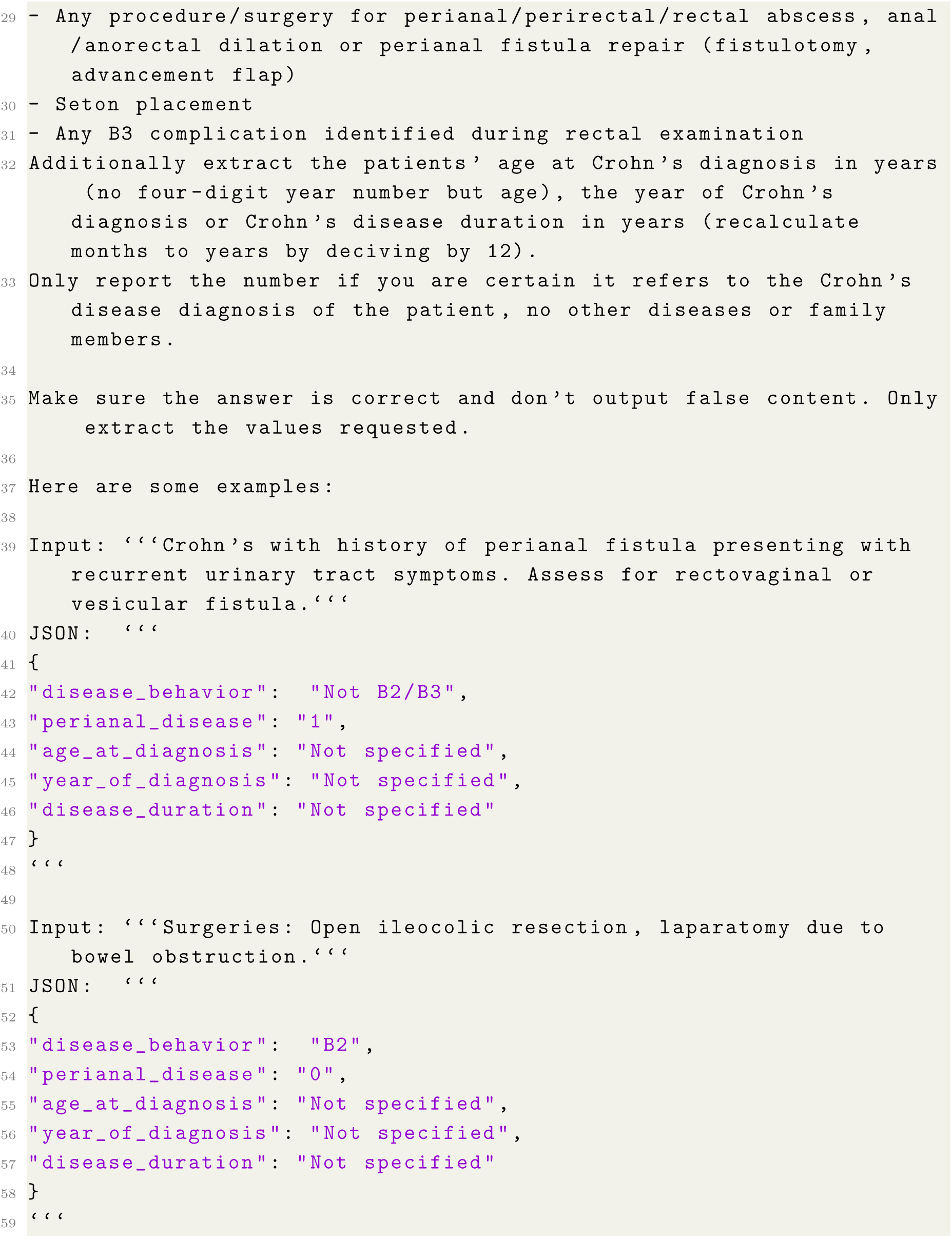
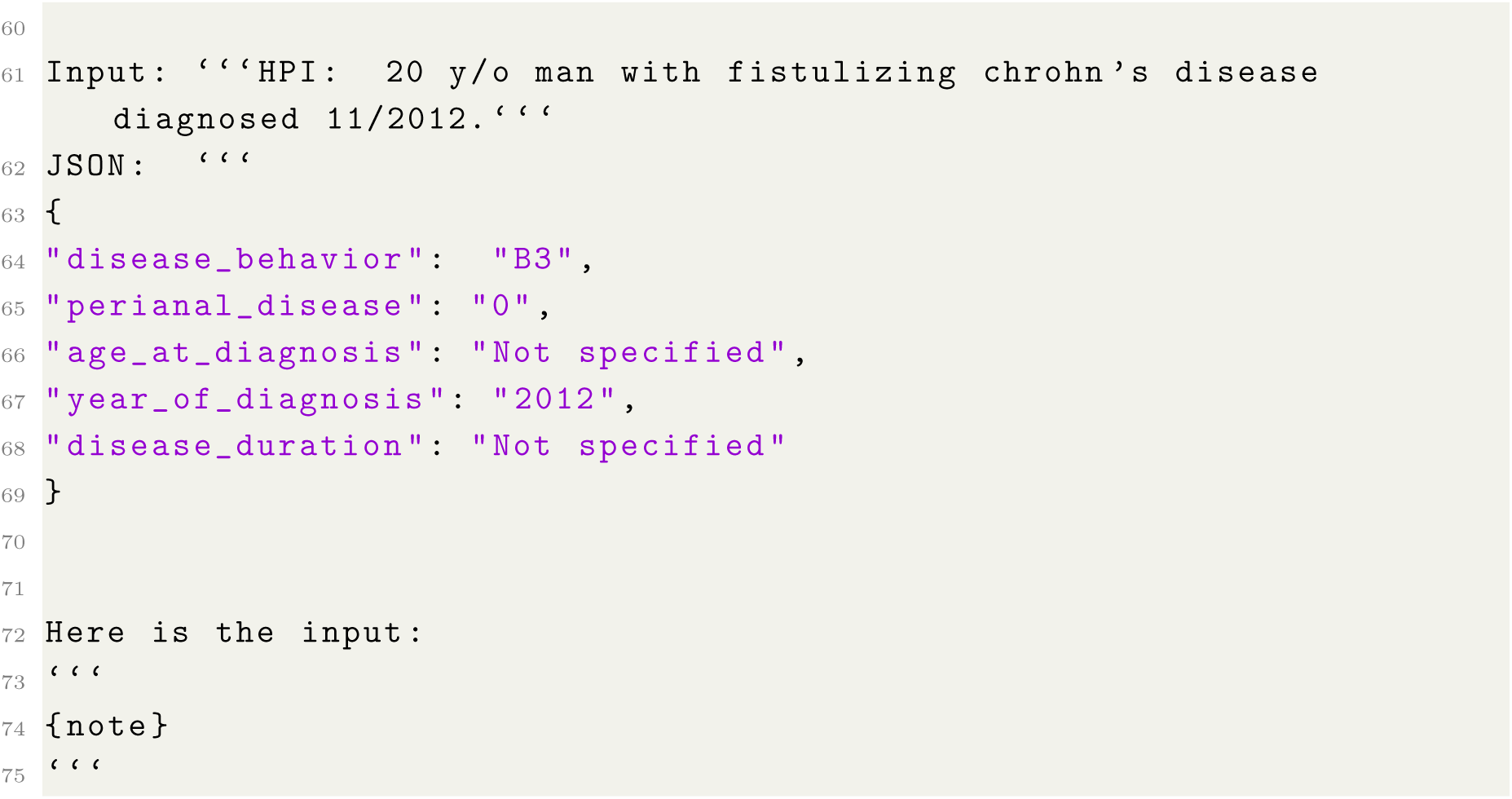
Prompt for GPT4-based phenotyping of disease behavior and age at diagnosis. Examples were randomly drawn from the labeled development dataset.

**Table A2:** Overview of the annotation process of the test dataset for the behavioral disease phenotype using clinical notes and radiology reports.

|  |  | Clinical Notes | Radiology Reports | Total |
| --- | --- | --- | --- | --- |
| **Total number of notes** |  | 150 | 50 | 200 |
| **Total number of sentences** |  | 14,236 | 1,154 | 15,390 |
| **Mean sentences per note (SD)** | | 95 ($\pm 87$) | 23 ($\pm 11$) | - |
| **Not B2/B3** | **Notes** | 112 | 32 | 144 |
|  | **Sentences** | 14,094 | 1,113 | 15,207 |
| **B2** | **Notes** | 13 | 7 | 20 |
|  | **Sentences** | 62 | 24 | 86 |
| **B3** | **Notes** | 25 | 11 | 36 |
|  | **Sentences** | 80 | 17 | 97 |
| **Perianal disease** | **Notes** | 25 | 7 | 32 |
|  | **Sentences** | 113 | 24 | 137 |

**Table A3:** Cohen’s kappa agreement scores calculated as inter-annotator agreements (IAA) between the two annotators on sentence- and note-level, as well as between the consensus labels from the two annotators, and the labels derived using GPT-4 or the rules on note-level, respectively.

| Comparison | Not B2/B3 | B2/B3 | Perianal | Average |
| --- | --- | --- | --- | --- |
| Sentence-level IAA | 0.83 | 0.84 | 0.87 | 0.85 |
| Note-level IAA | 0.88 | 0.88 | 0.95 | 0.90 |
| GPT-4/consensus IAA | 0.80 | 0.80 | 0.89 | 0.83 |
| Rules/consensus IAA | 0.84 | 0.84 | 0.90 | 0.86 |

**Table A4:** Overview of the annotation process of the test dataset for the age at diagnosis using clinical notes and radiology reports.

|  |  | Clinical Notes | Radiology Reports | Total |
| --- | --- | --- | --- | --- |
| **Total number of notes** |  | 79 | 1 | 80 |
| **Total number of sentences** |  | 12,261 | 32 | 12,293 |
| **Mean sentences per note (SD)** | | 155 ($\pm 116$) | 32 (NA) | - |
| **Age at diagnosis** | **Notes** | 18 | 0 | 18 |
|  | **Sentences** | 21 | 0 | 21 |
| **Year of diagnosis** | **Notes** | 47 | 1 | 48 |
|  | **Sentences** | 76 | 1 | 77 |
| **Disease duration** | **Notes** | 8 | 0 | 8 |
|  | **Sentences** | 10 | 0 | 10 |

**Table A5:** Performance of the rule-based phenotyping algorithm and GPT-4-based results of disease behavior phenotyping on note level using the newly annotated test dataset comprising in total 200 clinical notes and 50 radiology reports.

| Model | Phenotype | Recall | Precision | F1 score | Specificity |
| --- | --- | --- | --- | --- | --- |
| Rule-based | Not B2/B3 | 0.94 | 1.00 | 0.97 | 1.00 |
| Rule-based | B2 | 0.92 | 0.75 | 0.83 | 0.97 |
| Rule-based | B3 | 1.00 | 0.86 | 0.93 | 0.97 |
| GPT-4 | Not B2/B3 | 0.95 | 0.98 | 0.96 | 0.95 |
| GPT-4 | B2 | 0.92 | 0.57 | 0.71 | 0.93 |
| GPT-4 | B3 | 0.80 | 0.95 | 0.87 | 0.99 |

**Table A6:** Statistical differences between the Cohen’s kappa agreement scores from the note-level inter-annotator agreement (IAA) of the two annotators, as well as between the consensus labels from the two annotators, and the labels derived using GPT-4 or the rules on note-level, respectively. Differences are reported in form of the Z-statistic and the corresponding p-values and q-values (FDR-adjusted p-values).

| Phenotype | GPT-4 vs note-level IAA |  |  | Rules vs note-level IAA |  |  |
| --- | --- | --- | --- | --- | --- | --- |
|  | Z-score | p-value | q-value | Z-score | p-value | q-value |
| Not B2/B3 | -0.74 | 0.46 | 0.70 | -0.39 | 0.70 | 0.70 |
| B2/B3 | -0.74 | 0.46 | 0.70 | -0.39 | 0.70 | 0.70 |
| Perianal | -0.78 | 0.43 | 0.70 | -0.67 | 0.50 | 0.70 |

**Table A7:**
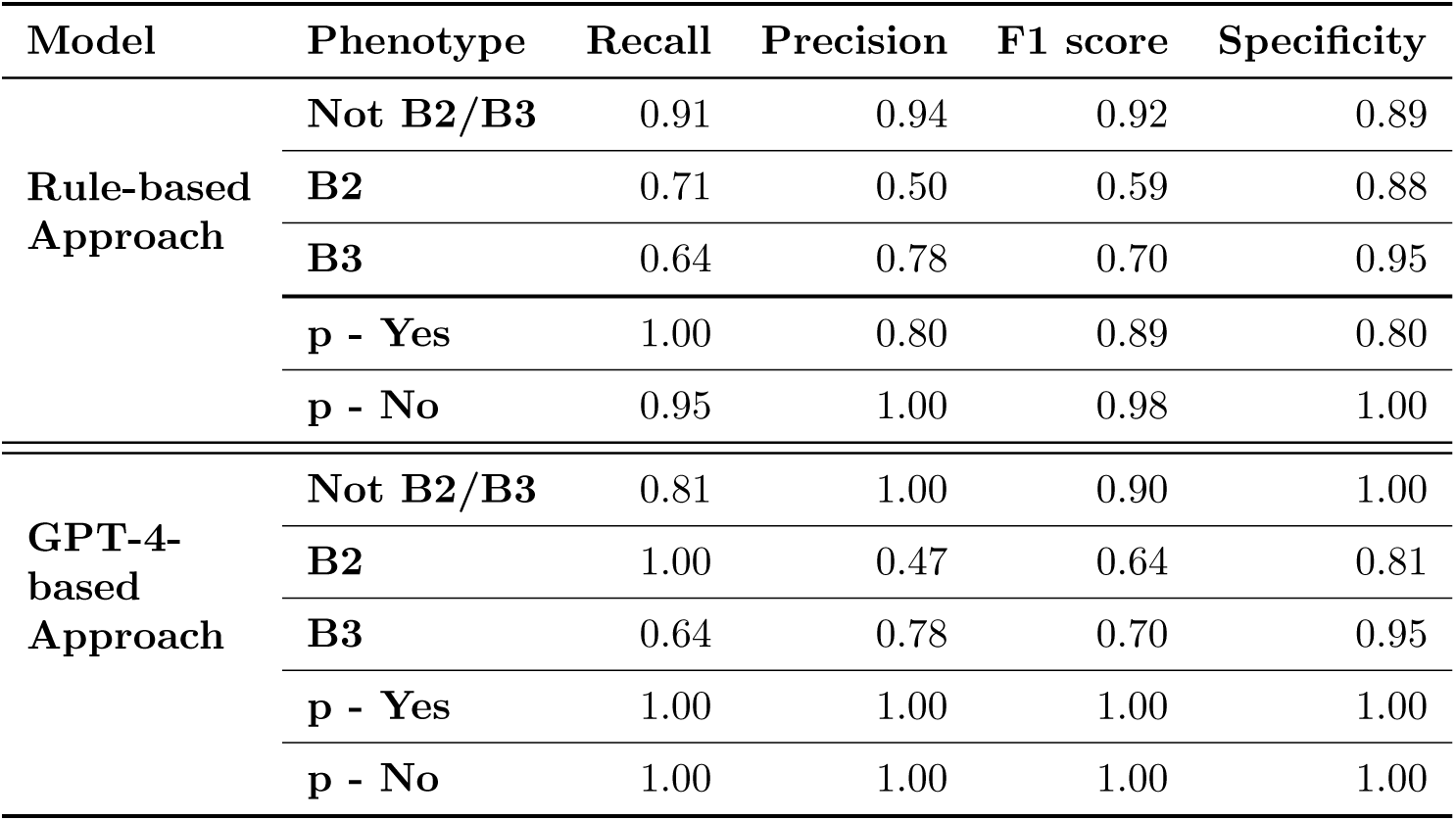
Performance of the rule-based phenotyping algorithm and GPT-4-based results of disease behavior phenotyping on note level using the newly annotated test dataset comprising 50 radiology reports.

| Model | Phenotype | Recall | Precision | F1 score | Specificity |
| --- | --- | --- | --- | --- | --- |
| Rule-based Approach | Not B2/B3 | 0.91 | 0.94 | 0.92 | 0.89 |
|  | B2 | 0.71 | 0.50 | 0.59 | 0.88 |
|  | B3 | 0.64 | 0.78 | 0.70 | 0.95 |
|  | p - Yes | 1.00 | 0.80 | 0.89 | 0.80 |
|  | p - No | 0.95 | 1.00 | 0.98 | 1.00 |
| GPT-4-based Approach | Not B2/B3 | 0.81 | 1.00 | 0.90 | 1.00 |
|  | B2 | 1.00 | 0.47 | 0.64 | 0.81 |
|  | B3 | 0.64 | 0.78 | 0.70 | 0.95 |
|  | p - Yes | 1.00 | 1.00 | 1.00 | 1.00 |
|  | p - No | 1.00 | 1.00 | 1.00 | 1.00 |

**Table A8:**
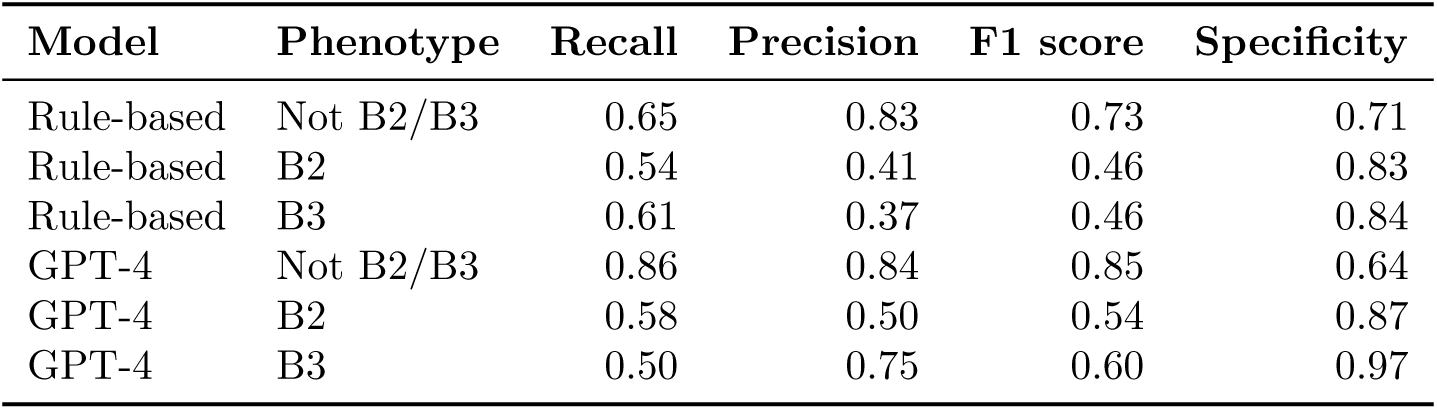
Performance of the rule-based phenotyping algorithm and GPT-4-based results of disease behavior phenotyping on patient level. 134 patients of the MSCCR cohort had available information on the behavioral disease phenotype through manual chart review.

| **Model** | **Phenotype** | **Recall** | **Precision** | **F1 score** | **Specificity** |
| --- | --- | --- | --- | --- | --- |
| Rule-based | Not B2/B3 | 0.65 | 0.83 | 0.73 | 0.71 |
| Rule-based | B2 | 0.54 | 0.41 | 0.46 | 0.83 |
| Rule-based | B3 | 0.61 | 0.37 | 0.46 | 0.84 |
| GPT-4 | Not B2/B3 | 0.86 | 0.84 | 0.85 | 0.64 |
| GPT-4 | B2 | 0.58 | 0.50 | 0.54 | 0.87 |
| GPT-4 | B3 | 0.50 | 0.75 | 0.60 | 0.97 |

## Supplementary Figures

**Figure A1:**
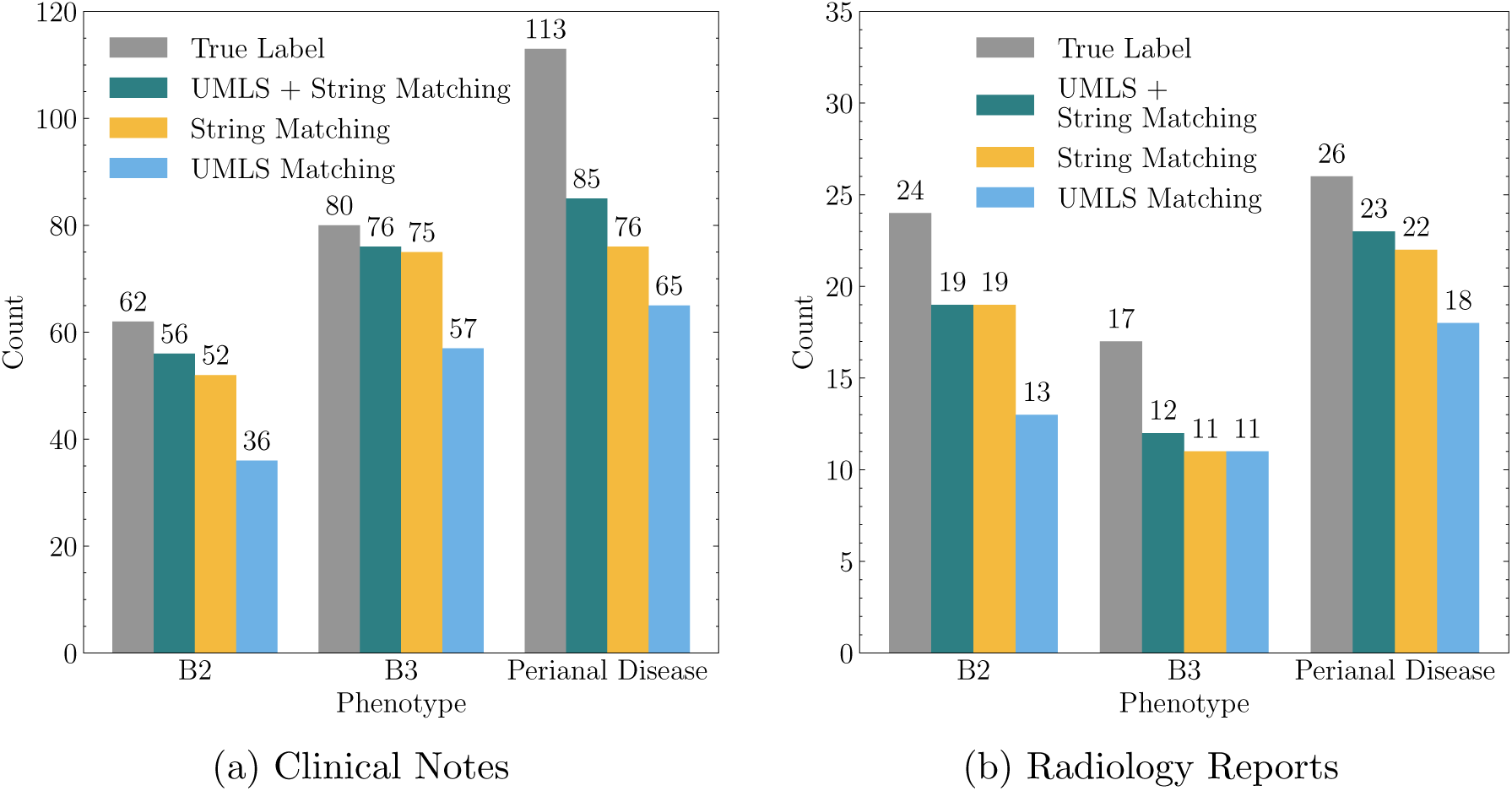
Correctly identified phenotypes by patterns. The count of correctly identified phenotypes by different usage of patterns versus the true count per phenotype on sentence level in the annotated clinical notes and radiology reports. UMLS matching refers to patterns using the matched UMLS codes, while string matching refers to patterns manually created to match specific phenotypes.

**Figure A2:**
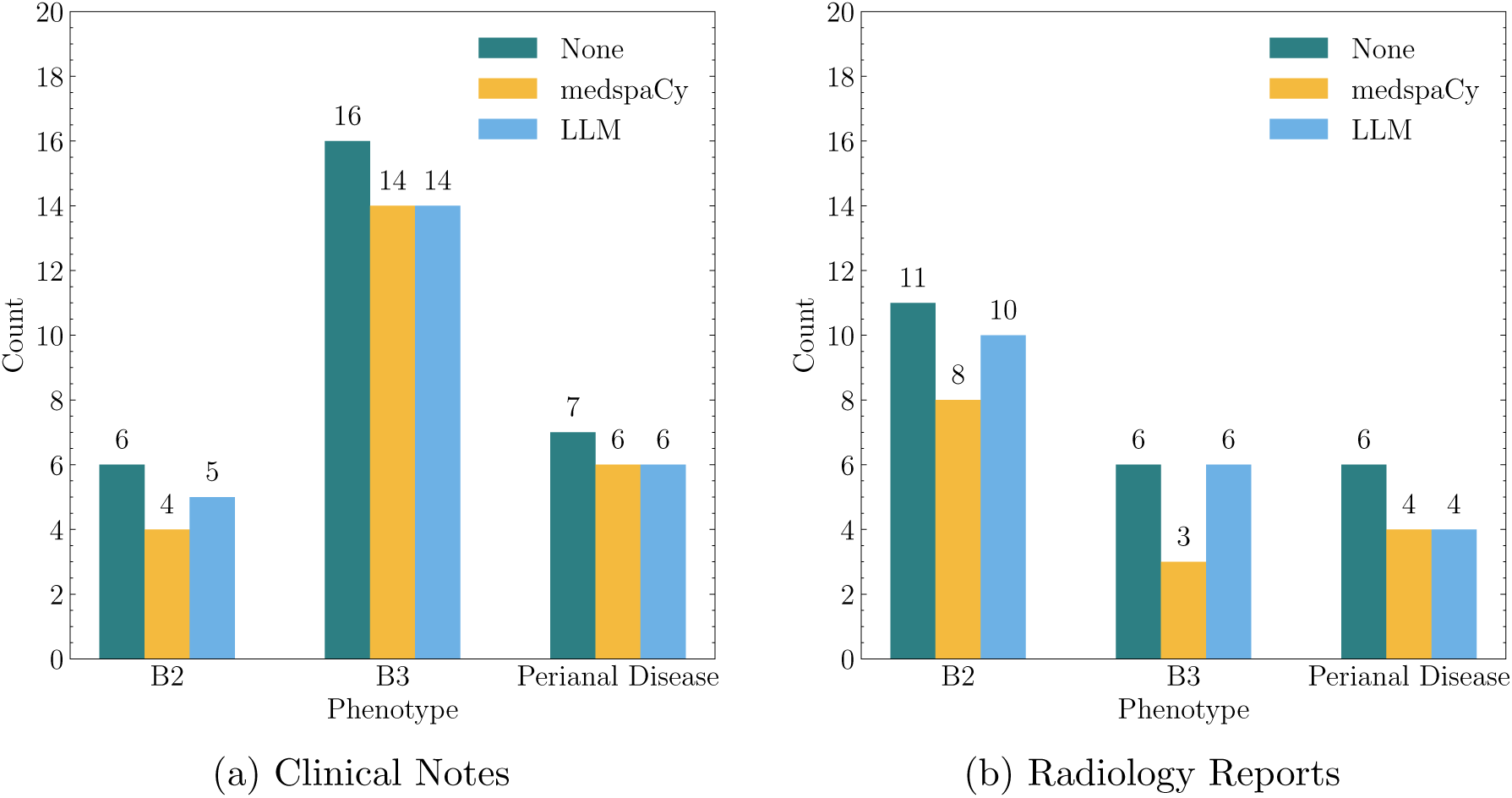
Incorrect identified phenotypes by negation detection methods. Differences in negation detection methods when analyzing the count of incorrectly identified phenotypes in the annotated clinical notes and radiology reports. “None” means, that no additional negation detection besides the manually defined rules for uncertainty and exclusion is used. “medspaCy” refers to negation detection based on the medspaCy ConText component, and “LLM” refers to negation detection via the clinical-assertion-negation-bert classifier.

**Figure A3:**
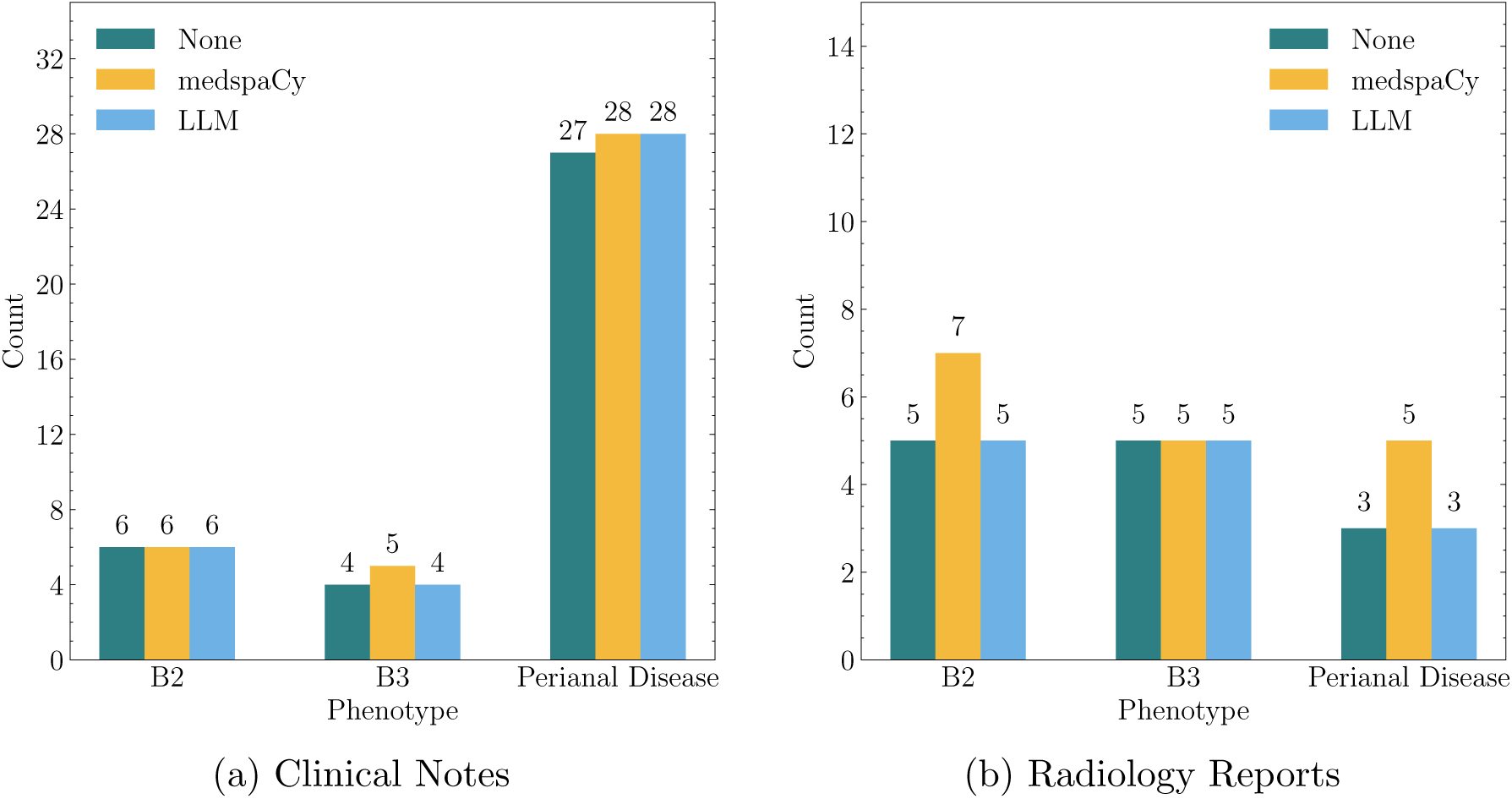
False negative phenotypes by negation detection methods. Differences in negation detection methods when analyzing the count of false negative phenotypes in the annotated clinical notes and radiology reports. “None” means, that no additional negation detection besides the manually defined rules for uncertainty and exclusion is used. “medspaCy” refers to negation detection based on the medspaCy ConText component, and “LLM” refers to negation detection via the clinical-assertion-negationbert classifier.

**Figure A4:**
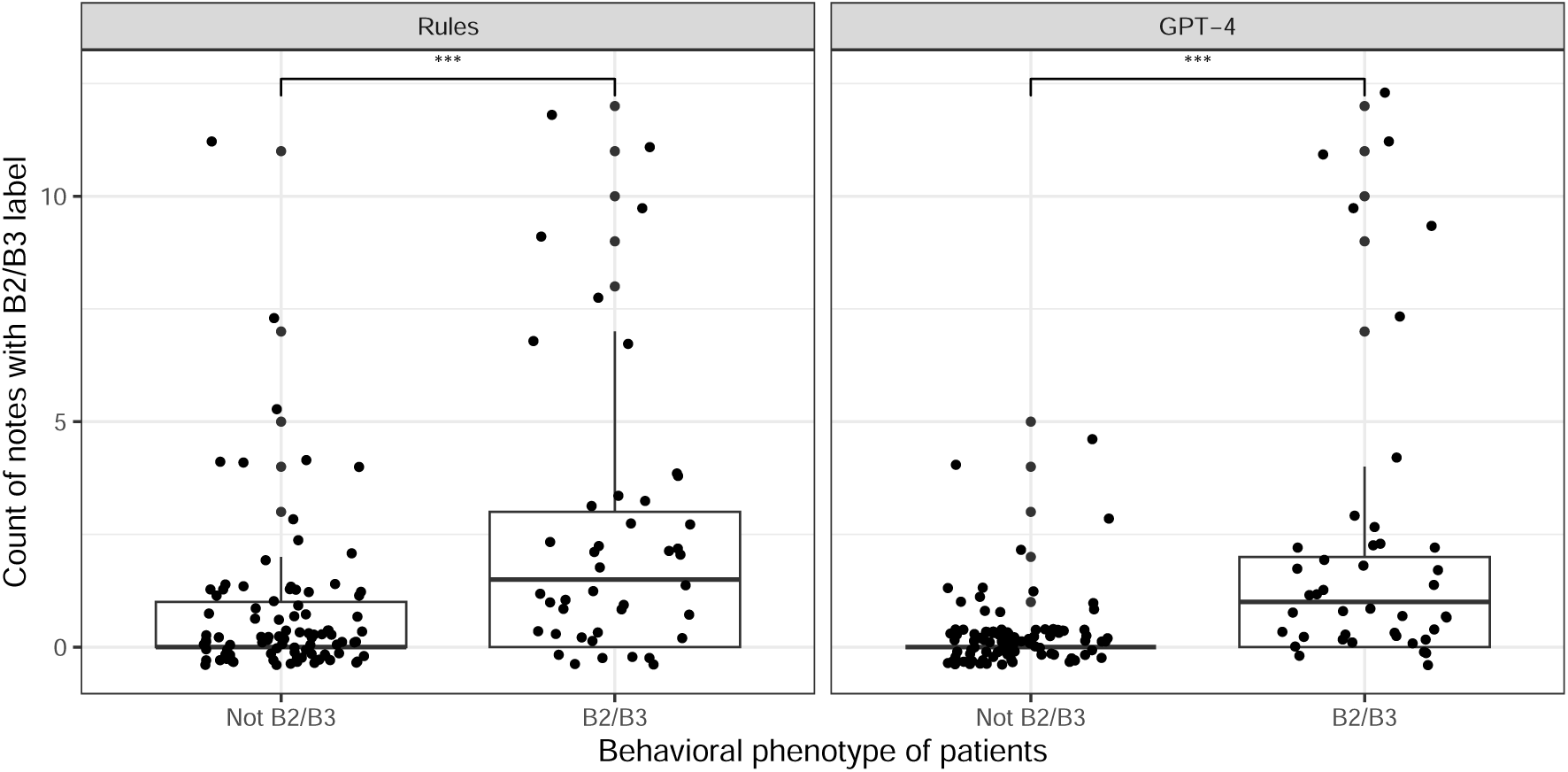
Counts of as penetrating disease classified notes per patient and phenotype group. 24 of the 134 labeled patients had stricturing disease (B2) and 18 penetrating disease (B3) at time point of study enrollment. Difference in note count between the groups was calculated using the Wilcoxon rank-sum test. NS.: not significant; “*”: p-value *<* 0.05; “**”: p-value *<* 0.01; “***”: p-value *<* 0.001.

**Figure A5:**
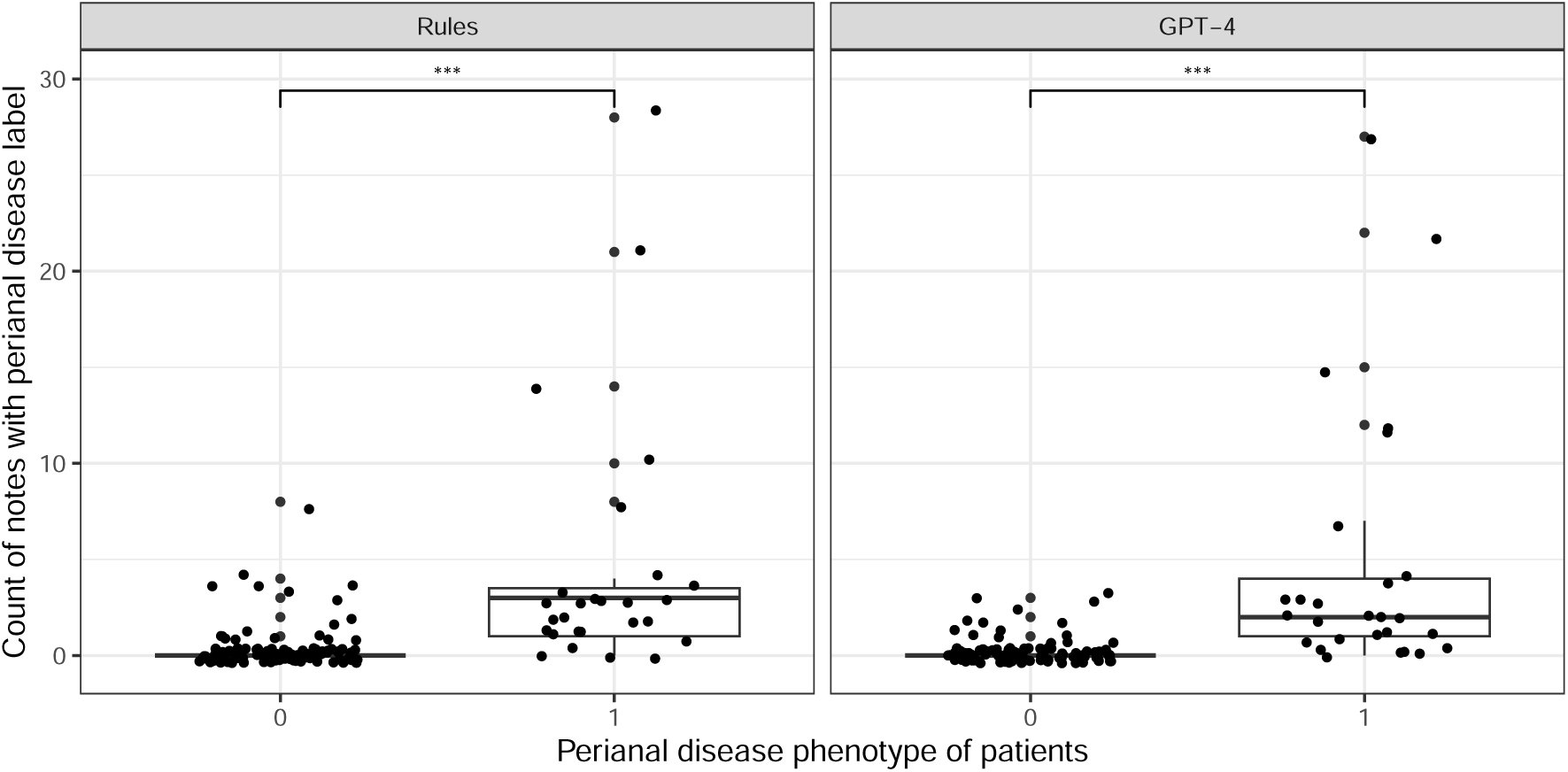
Counts of as perianal disease classified notes per patient and phenotype group. 27 of the 134 labeled patients had perianal disease at time point of study enrollment. Difference in note count between the groups was calculated using the Wilcoxon rank-sum test. NS.: not significant; “*”: p-value *<* 0.05; “**”: p-value *<* 0.01; “***”: p-value *<* 0.001.

**Figure A6:**
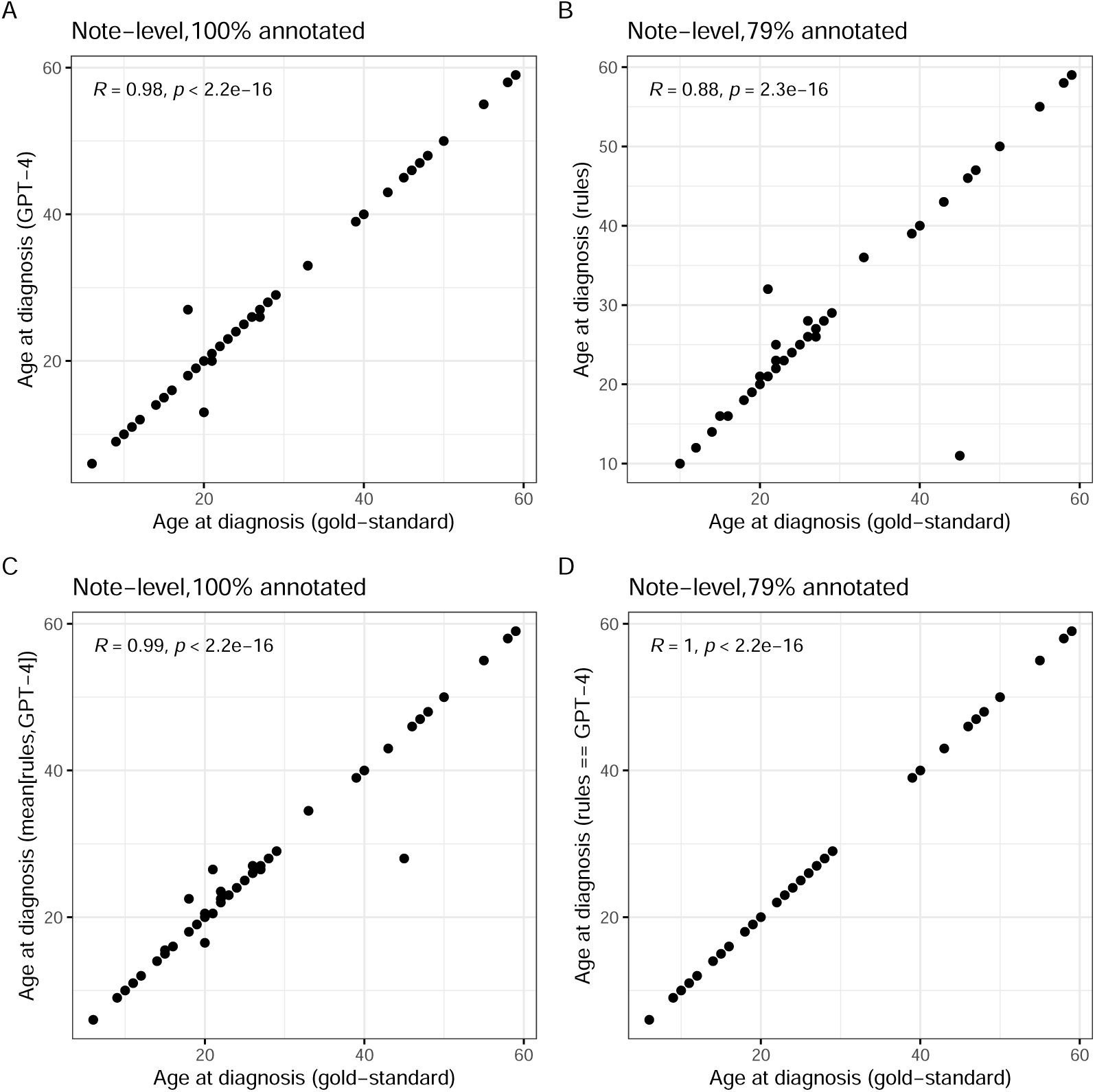
Pearson correlation of ground-truth age at diagnosis derived manual annotation and automatically extracted age at diagnosis values on note-level. Automatic extraction of age at diagnosis included (A) a GPT-4-based approach, (B) a rule-based approach, (C) a combination of the two, choosing the mean age at diagnosis value, and (D) the subset of notes where the extracted age at diagnosis value through rules and GPT-4 was identical. Fraction of notes that were annotated by each approach given in the header title.

**Figure A7:**
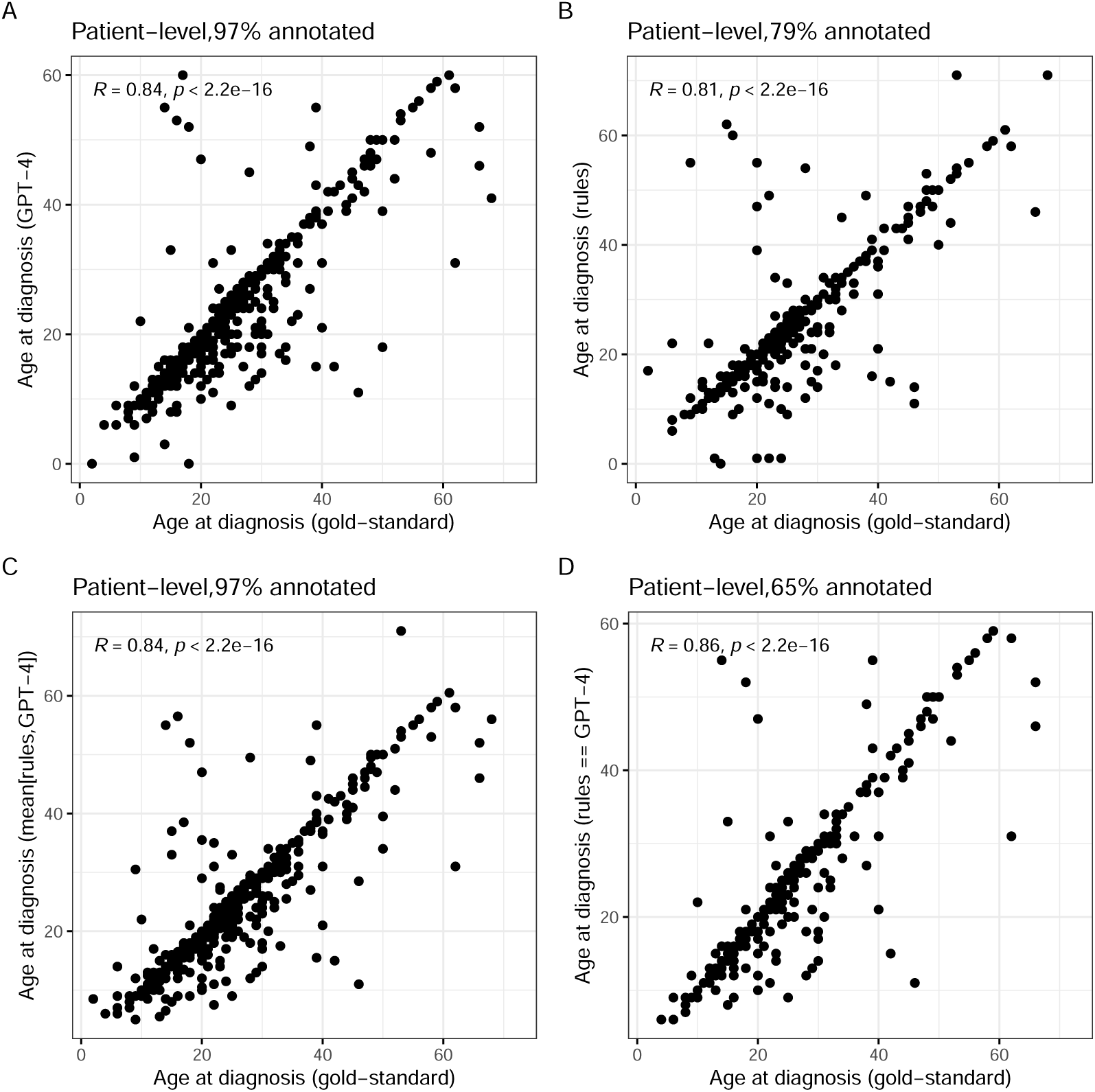
Pearson correlation of ground-truth age at diagnosis derived manual annotation and automatically extracted age at diagnosis values on patient-level. Automatic extraction of age at diagnosis included (A) a GPT-4-based approach, (B) a rule-based approach, (C) a combination of the two, choosing the mean age at diagnosis value, and (D) the subset of notes where the extracted age at diagnosis value through rules and GPT-4 was identical. Fraction of patients that were annotated by each approach given in the header title.

## Footnotes

1 Included clinical texts needed to match at least one of the following patterns: (D|d)iagnosed|DIAGNOSED, ((C|c)rohn|CROHN|cd|CD)[^a-zA-Z0-9]*(since|SINCE), (D|d)isease[^a-zA-Z0-9]*(O|o)nset|DISEASE[^a-zA-Z0-9]*ONSET, (A|a)ge[^a-zA-Z0-9]*(A|a)t[^a-zA-Z0-9]*(D|d)iagnosis, AGE[^a-zA-Z0-9]*AT[^a-zA-Z0-9]*DIAGNOSIS

2 https://huggingface.co/bvanaken/clinical-assertion-negation-bert

## References

[1] C. Shivade, P. Raghavan, E. Fosler-Lussier, et al., “A review of approaches to identifying patient phenotype cohorts using electronic health records,” Journal of the American Medical Informatics Association, vol. 21, no. 2, pp. 221–230, Mar. 2014, issn: 1067-5027. doi: 10.1136/amiajnl-2013-001935. [Online]. Available: 10.1136/amiajnl-2013-001935 (visited on 07/20/2024).

[2] T. He, A. Belouali, J. Patricoski, et al., “Trends and opportunities in computable clinical phenotyping: A scoping review,” Journal of Biomedical Informatics, vol. 140, p. 104 335, Apr. 2023, issn: 1532-0464. doi: 10.1016/j.jbi.2023.104335. [Online]. Available: https://www.sciencedirect.com/science/article/pii/S1532046423000564 (visited on 07/21/2024).

[3] J. C. Kirby, P. Speltz, L. V. Rasmussen, et al., “PheKB: A catalog and workflow for creating electronic phenotype algorithms for transportability,” eng, Journal of the American Medical Informatics Association: JAMIA, vol. 23, no. 6, pp. 1046–1052, Nov. 2016, issn: 1527-974X. doi: 10.1093/jamia/ocv202.

[4] B. Khosravi, P. Rouzrokh, and B. J. Erickson, “Getting More Out of Large Databases and EHRs with Natural Language Processing and Artificial Intelligence: The Future Is Here,” en-US, JBJS, vol. 104, no. Suppl 3, p. 51, Oct. 2022, issn: 0021-9355. doi: 10.2106/JBJS.22.00567.

[5] Z. Zeng, Y. Deng, X. Li, T. Naumann, and Y. Luo, “Natural Language Processing for EHR-Based Computational Phenotyping,” IEEE/ACM Transactions on Computational Biology and Bioinformatics, vol. 16, no. 1, pp. 139–153, Jan. 2019, Conference Name: IEEE/ACM Transactions on Computational Biology and Bioinformatics, issn: 1557-9964. doi: 10.1109/TCBB.2018.2849968.

[6] A. Moldwin, D. Demner-Fushman, and T. R. Goodwin, “Empirical Findings on the Role of Structured Data, Unstructured Data, and their Combination for Automatic Clinical Phenotyping,” AMIA Summits on Translational Science Proceedings, vol. 2021, pp. 445–454, May 2021, issn: 2153-4063.

[7] A. N. Ananthakrishnan, T. Cai, G. Savova, et al., “Improving Case Definition of Crohn’s Disease and Ulcerative Colitis in Electronic Medical Records Using Natural Language Processing: A Novel Informatics Approach,” Inflammatory bowel diseases, vol. 19, no. 7, pp. 1411–1420, Jun. 2013, issn: 1078-0998. doi: 10.1097/MIB.0b013e31828133fd.

[8] J. Torres, S. Mehandru, J.-F. Colombel, and L. Peyrin-Biroulet, “Crohn’s disease,” en, The Lancet, vol. 389, no. 10080, pp. 1741–1755, Apr. 2017, issn: 0140-6736. doi: 10.1016/S0140-6736(16)31711-1.

[9] J. Hanzel, P. Bossuyt, V. Pittet, et al., “Development of a Core Outcome Set for Real-world Data in Inflammatory Bowel Disease: A European Crohn’s and Colitis Organisation [ECCO] Position Paper,” Journal of Crohn’s and Colitis, jjac136, Oct. 2022, issn: 1873-9946. doi: 10.1093/ecco-jcc/jjac136.

[10] L. Blonde, K. Khunti, S. B. Harris, C. Meizinger, and N. S. Skolnik, “Interpretation and Impact of Real-World Clinical Data for the Practicing Clinician,” Advances in Therapy, vol. 35, no. 11, pp. 1763–1774, 2018, issn: 0741-238X. doi: 10.1007/s12325-018-0805-y. [Online]. Available: https://www.ncbi.nlm.nih.gov/pmc/articles/PMC6223979/ (visited on 10/11/2023).

[11] C. Ha, T. A. Ullman, C. A. Siegel, and A. Kornbluth, “Patients Enrolled in Randomized Controlled Trials Do Not Represent the Inflammatory Bowel Disease Patient Population,” Clinical Gastroenterology and Hepatology, vol. 10, no. 9, pp. 1002–1007, Sep. 2012, issn: 1542-3565. doi: 10.1016/j.cgh.2012.02.004. [Online]. Available: https://www.sciencedirect.com/science/article/pii/S1542356512002017 (visited on 10/11/2023).

[12] M. S. Silverberg, J. Satsangi, T. Ahmad, et al., “Toward an integrated clinical, molecular and serological classification of inflammatory bowel disease: Report of a Working Party of the 2005 Montreal World Congress of Gastroenterology,” eng, Canadian Journal of Gastroenterology = Journal Canadien De Gastroenterologie, vol. 19 Suppl A, 5A–36A, Sep. 2005, issn: 1916-7237. doi: 10.1155/2005/269076.

[13] J. Satsangi, M. S. Silverberg, S. Vermeire, and J.-F. Colombel, “The Montreal classification of inflammatory bowel disease: Controversies, consensus, and implications,” en, Gut, vol. 55, no. 6, pp. 749–753, Jun. 2006, Publisher: BMJ Publishing Group Section: Leading article, issn: 0017-5749, 1468-3288. doi: 10.1136/gut.2005.082909.

[14] F. Rieder, E. M. Zimmermann, F. H. Remzi, and W. J. Sandborn, “Crohn’s disease complicated by strictures: A systematic review,” en, Gut, vol. 62, no. 7, pp. 1072–1084, Jul. 2013, Publisher: BMJ Publishing Group Section: Recent advances in clinical practice, issn: 0017-5749, 1468-3288. doi: 10.1136/gutjnl-2012-304353.

[15] M. Scharl and G. Rogler, “Pathophysiology of fistula formation in Crohn’s disease,” World Journal of Gastrointestinal Pathophysiology, vol. 5, no. 3, pp. 205–212, Aug. 2014, issn: 2150-5330. doi: 10.4291/wjgp.v5.i3.205.

[16] A. T. P. Carvalho, B. C. Esberard, and A. da Luz Moreira, “Current management of spontaneous intra-abdominal abscess in Crohn’s disease,” en, Journal of Coloproctology, vol. 38, no. 2, pp. 158–163, Apr. 2018, issn: 2237-9363. doi: 10.1016/j.jcol.2016.05.003.

[17] B. Pariente, J.-Y. Mary, S. Danese, et al., “Development of the Lémann index to assess digestive tract damage in patients with Crohn’s disease,” eng, Gastroenterology, vol. 148, no. 1, 52–63.e3, Jan. 2015, issn: 1528-0012. doi: 10.1053/j.gastro.2014.09.015.

[18] R. Kosoy, S. Kim-Schulze, A. Rahman, et al., “Deep Analysis of the Peripheral Immune System in IBD Reveals New Insight in Disease Subtyping and Response to Monotherapy or Combination Therapy,” en, Cellular and Molecular Gastroenterology and Hepatology, vol. 12, no. 2, pp. 599–632, Jan. 2021, issn: 2352-345X. doi: 10.1016/j.jcmgh.2021.03.012.

[19] S. L. Gold, L. G. Rabinowitz, L. Manning, et al., “High Prevalence of Malnutrition and Micronutrient Deficiencies in Patients With Inflammatory Bowel Disease Early in Disease Course,” Inflammatory Bowel Diseases, vol. 29, no. 3, pp. 423–429, May 2022, issn: 1078-0998. doi: 10.1093/ibd/izac102.

[20] B. E. Sands, N. LeLeiko, S. A. Shah, R. Bright, and S. Grabert, “OSCCAR: Ocean State Crohn’s and Colitis Area Registry,” eng, Medicine and Health, Rhode Island, vol. 92, no. 3, pp. 82–85, 88, Mar. 2009, issn: 1086-5462.

[21] S. Ben-Horin, L. Novack, R. Mao, et al., “Efficacy of Biologic Drugs in Short-Duration Versus Long-Duration Inflammatory Bowel Disease: A Systematic Review and an Individual-Patient Data Meta-Analysis of Randomized Controlled Trials,” Gastroenterology, vol. 162, no. 2, pp. 482–494, 2022, Publisher: The Authors, issn: 15280012. doi: 10.1053/j.gastro.2021.10.037. [Online]. Available: 10.1053/j.gastro.2021.10.037.

[22] R. W. Stidham, D. Yu, X. Zhao, et al., “Identifying the Presence, Activity, and Status of Extraintestinal Manifestations of Inflammatory Bowel Disease Using Natural Language Processing of Clinical Notes,” Inflammatory Bowel Diseases, vol. 29, no. 4, pp. 503–510, Apr. 2023, issn: 1078-0998. doi: 10.1093/ibd/izac109. [Online]. Available: 10.1093/ibd/izac109 (visited on 10/11/2023).

[23] Icahn School of Medicine at Mount Sinai, Mount Sinai Data Warehouse — Scientific Computing and Data, en-US, 2023. [Online]. Available: https://labs.icahn.mssm.edu/msdw/ (visited on 07/11/2023).

[24] J. Cohen, “A Coefficient of Agreement for Nominal Scales,” en, Educational and Psychological Measurement, vol. 20, no. 1, pp. 37–46, Apr. 1960, issn: 0013-1644, 1552-3888. doi: 10.1177/001316446002000104.

[25] M. Honnibal, I. Montani, S. Van Landeghem, and A. Boyd, spaCy: Industrial-strength Natural Language Processing in Python, original-date: 2014-07-03, 2020. doi: 10.5281/zenodo.1212303. [Online]. Available: https://github.com/explosion/spaCy (visited on 06/10/2023).

[26] M. Neumann, D. King, I. Beltagy, and W. Ammar, “ScispaCy: Fast and Robust Models for Biomedical Natural Language Processing,” en, Proceedings of the 18th BioNLP Workshop and Shared Task, pp. 319–327, 2019, Conference Name: Proceedings of the 18th BioNLP Workshop and Shared Task Place: Florence, Italy Publisher: Association for Computational Linguistics. doi: 10.18653/v1/W19-5034.

[27] H. Eyre, A. B. Chapman, K. S. Peterson, et al., Launching into clinical space with medspaCy: A new clinical text processing toolkit in Python, arXiv:2106.07799 [cs], Jun. 2021. doi: 10.48550/arXiv.2106.07799.

[28] B. van Aken, I. Trajanovska, A. Siu, M. Mayrdorfer, K. Budde, and A. Loeser, “Assertion Detection in Clinical Notes: Medical Language Models to the Rescue?” In Proceedings of the Second Workshop on Natural Language Processing for Medical Conversations, Online: Association for Computational Linguistics, Jun. 2021, pp. 35–40. doi: 10.18653/v1/2021.nlpmc-1.5.

[29] OpenAI, J. Achiam, S. Adler, et al., GPT-4 Technical Report, arXiv:2303.08774 [cs], Mar. 2024. doi: 10.48550/arXiv.2303.08774. [Online]. Available: http://arxiv.org/abs/2303.08774 (visited on 07/25/2024).

[30] J. R. Landis and G. G. Koch, “The measurement of observer agreement for categorical data,” eng, Biometrics, vol. 33, no. 1, pp. 159–174, Mar. 1977, issn: 0006-341X.

[31] M. C. V. Lieke M Spekhorst and D. I. o. C. a. Colitis (ICC), “Performance of the Montreal classification for inflammatory bowel diseases,” en, World Journal of Gastroenterology, vol. 20, no. 41, pp. 15 374–15 381, Nov. 2014, Publisher: Baishideng Publishing Group Inc. doi: 10.3748/wjg.v20.i41.15374.

[32] K. Krishnaprasad, J. M. Andrews, I. C. Lawrance, et al., “Inter-observer agreement for Crohn’s disease sub-phenotypes using the Montreal Classification: How good are we? A multi-centre Australasian study,” Journal of Crohn’s and Colitis, vol. 6, no. 3, pp. 287–293, Apr. 2012, issn: 1873-9946. doi: 10.1016/j.crohns.2011.08.016.

[33] T. Dassopoulos, G. C. Nguyen, A. Bitton, et al., “Assessment of reliability and validity of IBD phenotyping within the National Institutes of Diabetes and Digestive and Kidney Diseases (NIDDK) IBD Genetics Consortium (IBDGC),” Inflammatory Bowel Diseases, vol. 13, no. 8, pp. 975–983, Aug. 2007, issn: 1078-0998. doi: 10.1002/ibd.20144. [Online]. Available: 10.1002/ibd.20144 (visited on 07/25/2024).

[34] S. Shrestha, O. Olén, C. Eriksson, et al., “The use of ICD codes to identify IBD subtypes and phenotypes of the Montreal classification in the Swedish National Patient Register,” eng, Scandinavian Journal of Gastroenterology, vol. 55, no. 4, pp. 430–435, Apr. 2020, issn: 1502-7708. doi: 10.1080/00365521.2020.1740778.

[35] A. N. Kho, J. Yu, M. S. Bryan, et al., “Privacy-Preserving Record Linkage to Identify Fragmented Electronic Medical Records in the All of Us Research Program,” en, in Machine Learning and Knowledge Discovery in Databases, P. Cellier and K. Driessens, Eds., ser. Communications in Computer and Information Science, Cham: Springer International Publishing, 2020, pp. 79–87, isbn: 978-3-030-43887-6. doi: 10.1007/978-3-030-43887-6_7.

[36] X. Luo, F. M. Tahabi, T. Marc, L. A. Haunert, and S. Storey, “Zero-shot learning to extract assessment criteria and medical services from the preventive healthcare guidelines using large language models,” Journal of the American Medical Informatics Association, vol. 31, no. 8, pp. 1743–1753, Aug. 2024, issn: 1527-974X. doi: 10.1093/jamia/ocae145. [Online]. Available: 10.1093/jamia/ocae145 (visited on 08/27/2024).

[37] Z. Chen, A. H. Cano, A. Romanou, et al., MEDITRON-70B: Scaling Medical Pretraining for Large Language Models, arXiv:2311.16079 [cs], Nov. 2023. doi: 10.48550/arXiv.2311.16079. [Online]. Available: http://arxiv.org/abs/2311.16079 (visited on 07/22/2024).

[38] Y. Labrak, A. Bazoge, E. Morin, P.-A. Gourraud, M. Rouvier, and R. Dufour, BioMistral: A Collection of Open-Source Pretrained Large Language Models for Medical Domains, arXiv:2402.10373 [cs], Jul. 2024. doi: 10.48550/arXiv.2402.10373. [Online]. Available: http://arxiv.org/abs/2402.10373 (visited on 07/22/2024).

